# Sex and hormonal effects on drug cue-reactivity and its regulation in human addiction

**DOI:** 10.1101/2024.11.18.24317491

**Authors:** Yuefeng Huang, Eduardo R. Butelman, Ahmet O. Ceceli, Greg Kronberg, Sarah G. King, Natalie E. McClain, Yui Ying Wong, Maggie Boros, K Rachel Drury, Rajita Sinha, Nelly Alia-Klein, Rita Z. Goldstein

**Author notes:** Corresponding author: Rita Z. Goldstein, Icahn School of Medicine at Mount Sinai Gustave L. Levy Place, Box 1230, New York, NY 10029.

## Abstract

**Objective:** To study the sex and hormonal effects on cortico-striatal engagement during drug cue-reactivity and its regulation focusing on drug reappraisal.

**Methods:** Forty-nine men (age=41.96±9.71) with heroin use disorder (HUD) and 32 age-matched women (age=38.85±9.84) with HUD (n=16) or cocaine use disorder (CUD; n=16) were scanned using functional MRI, with a subgroup of women scanned twice, during the late-follicular and mid-luteal phases, to examine sex and menstrual phase differences in cortico-striatal drug cue-reactivity and its cognitive reappraisal and their correlations with ovarian hormones and drug craving.

**Results:** Women showed higher medial prefrontal cortex (PFC) drug cue-reactivity while men showed higher frontal eye field (FEF)/dorsolateral PFC (dlPFC) drug reappraisal as associated with lower cue-induced drug craving. In the women, drug cue-reactivity was higher during the follicular phase in the FEF/dlPFC, whereas drug reappraisal was higher during the luteal phase in the anterior PFC/orbitofrontal cortex. The more the estradiol during the follicular vs. luteal phase (Δ), the higher the Δdrug cue-reactivity in the vmPFC, which also correlated with higher Δdrug craving (observed also in the inferior frontal gyrus). The more this Δestradiol, the lower the Δdrug reappraisal in the vmPFC, anterior PFC and striatum. Conversely, Δprogesterone/estradiol ratio was positively associated with Δdrug reappraisal in the dlPFC.

**Conclusions:** Compared to men, women with addiction show more cortico-striatal reactivity to drug cue exposure and less PFC activity during drug reappraisal, driven by the follicular compared to luteal phase and directly related to craving and fluctuations in estrogen and progesterone with the former constituting a vulnerability and the latter a protective factor. This study provides insights for developing precisely timed and hormonally informed treatments for women with addiction.

## Introduction

Women have been reported to exhibit greater severity of drug abuse, a faster progression to problematic drug use, and higher relapse risk than men (1–3) although these sex differences are not always robust, with opposite results also reported (4, 5). For example, even after controlling for overall levels of drug exposure, we recently reported that men have higher opioid, cocaine, and psychostimulant overdose mortality rates than women (6). These discrepancies may be partially driven by fluctuations in women in the two primary ovarian hormones (1–3), estradiol and progesterone, which have been suggested to exert opposing effects on addiction-related behaviors: estradiol facilitates the initiation of drug taking (1) and reinstatement of drug-seeking behavior (1, 5), while progesterone reduces craving (7), positive subjective drug effects (8), and amount (9) of substance use. Overall, the complex interplay of sex- and hormonal-based patterns of risk and resilience in addiction is understudied partly because women remain significantly underrepresented in basic neuroscience (including neuroimaging) studies (10, 11). Of particular relevance, merely five neuroimaging studies examined the neural correlates of sex differences in cue-induced reactivity and craving in cocaine use disorder (CUD) and none in heroin use disorder (HUD) as previously reviewed (5). Furthermore, studies of hormonal effects in women are scarce, particularly in substance use disorder (SUD) (12).

Enhanced salience of drug cues at the expense of non-drug reinforcers, associated with hyperactivity across reward, salience, and executive control networks (among others), is a hallmark of addiction (11). While limited in number, and although a mixed direction of effects across substances including cocaine and alcohol has been observed, clinical neuroimaging studies indicate consistent sex differences in ventromedial prefrontal cortex (vmPFC) drug cue-reactivity in people with SUD (13). For example, drug cue-reactivity in the vmPFC to alcohol pictures was found higher in women than men (14), correlate with alcohol craving, and predict heavy drinking days, with the latter exclusively in women (15). Importantly, ovarian hormones can influence drug cue-reactivity, whereby cigarette-dependent women display striatal hyperreactivity to smoking cues during the late-follicular phase (increased estradiol) as compared to the mid-luteal phase (increased progesterone) and early-follicular phase (low estradiol and progesterone) (12).

Given its association with prospective drug use and relapse (16), reducing drug cue-reactivity has been a main goal in addiction treatment (17). A commonly used behavioral intervention for this purpose is emotion downregulation via cognitive reappraisal of drug cues; emotion upregulation via savoring of non-drug alternative (reward) cues has also been used, albeit much less frequently (18, 19). In addition to the expected higher drug cue-reactivity in the vmPFC and striatum (and in the orbitofrontal cortex and inferior frontal gyrus [IFG]), we previously identified elevated cortico-striatal activity during drug reappraisal as directly compared to food savoring in individuals with HUD (iHUD). This activity in the dorsolateral PFC (dlPFC) correlated with higher methadone dose, while its direct contrast with drug cue-reactivity correlated with (lower) craving, suggesting that cognitive reappraisal is an effective, yet resource-demanding, regulation strategy (20). In the general population, studies found that men, compared to women, have higher dlPFC activation during emotion downregulation (>passive viewing) of negative images (21, 22). Additionally, although an opposite effect was reported in postmenopausal women with depression (23), in naturally cycling women, estradiol administration lowers IFG/middle frontal gyrus activity during emotion downregulation (24).

Progesterone has the opposite effect as suggested by a study where intrauterine release of a synthetic progestin (levonorgestrel) promoted higher frontal N2 amplitude during emotion upregulation of negative images, suggesting the recruitment of more attention and cognitive control resources (25, 26). These findings collectively suggest potential sex and hormonal effects also on the regulation of drug cue-reactivity with implications for women’s mental health.

Therefore, our study aimed to explore sex and hormonal effects on cortico-striatal mechanisms underlying drug cue-reactivity (vs. processing of non-drug reward and neutral cues) and its regulation in individuals with SUD. Given the limited and inconsistent findings, and especially absence of relevant studies in HUD, our first hypothesis posited significant sex differences in drug cue-reactivity, without specifying a direction for this effect. Reflecting risk and protective effects, respectively, we then hypothesized that women would exhibit higher drug cue-reactivity and lower drug reappraisal during the follicular phase, with the opposite pattern in the luteal phase, as directly correlated with fluctuations in estrogen, progesterone and craving. Due to the scarcity of studies on emotion regulation with alternative reward savoring, and space limitations, these results are reported in the Supplement.

## Methods

### Participants

Forty-nine men with HUD (age=41.96±9.71 and 32 age-matched women with HUD or CUD (iHUD: n=16; iCUD n=16; age=38.85±9.84) from the greater New York City area were recruited.

Participants underwent a comprehensive clinical diagnostic interview conducted by trained research staff under a clinical psychologist’s supervision. All iHUD and iCUD met DSM-5 criteria for opioid or stimulant use disorder (with heroin or cocaine as the primary drug of choice or reason for treatment, respectively). See supplement for details of interview procedures, exclusionary criteria, comorbidities, and drug use/treatment related information.

All participants were scanned with MRI once to examine sex differences. A subgroup of sixteen women (iCUD=13, iHUD=3) underwent an additional MRI procedure to examine menstrual cycle/hormone effects (days between scans=47.80±61.08). Menstrual phase information for both MRI days (follicular vs. luteal) was tracked via self-report and validated by objective blood ovarian hormone testing using chemiluminescence microparticle (Abbott Architect) or electrochemiluminescence (ECLIA, Roche) immunoassays (see Supplement). All 16 women had a regular menstrual cycle (past 3 months average days=30.52±2.21) and none used hormonal contraceptives (self-reported). They were randomized to have their first scan at the follicular (n=6) or luteal (n=10) phase, with no significant group differences in these numbers (p=0.289).

The study was approved by the IRB of the Icahn School of Medicine at Mount Sinai. All participants provided written informed consent. Throughout the manuscript, the terms "men" and "women" are used to describe self-reported biological sex.

### fMRI task paradigm

The task paradigm was reported in detail in our previous study (20). In short, participants were instructed to passively look at drug (images of drug preparation, use, and paraphernalia; heroin for iHUD, and cocaine for iCUD), food, and neutral images, actively downregulate their emotional reactivity to the drug images, and actively upregulate their emotional reactivity to food images, during the “look”, “reappraise”, and “savor” conditions, respectively. Immediately before and after the task, participants provided self-reported drug and food craving and task motivation ratings. Self-evaluations of the difficulty and effectiveness of reappraisal and savoring were also collected after the task. After the MRI session, each participant provided ratings on valence, arousal, and craving (cue-induced craving; for food and drug cues only) on half of the drug, food, and neutral images viewed during the task (see Supplement).

### BOLD fMRI data analysis

Individual parameter estimates were generated using the general linear model (GLM) via FMRIB Software Library (FSL)’s FEAT (version 6.0) (27). See Supplement and previous study (20) for GLM details. A fixed-effects model was used for subject-level statistical maps of each task event (look drug/food/neutral, reappraise drug, savor food) and their contrasts to yield estimates of drug cue-reactivity (look drug>look neutral or look food) and its regulation via reappraisal (reappraise drug>look drug) and savoring (savor food>look drug). For completeness, we also inspected the contrasts of reappraise drug>savor food and savor food>look food; the savoring results are reported in the Supplement. Group-level estimates were calculated using FSL FLAME 1+2 mixed-effects model to improve group-level variance estimation and population inferences via Markov chain Monte Carlo simulations (28). Given the nonuniformity in number of cue types per task condition, instead of calculating condition-related main effects and interactions, we preselected the above-mentioned contrasts of interest to test our hypotheses. For within-women (delta measures between menstrual phases) correlations, the delta directions are follicular-luteal phase for estradiol and drug craving and luteal-follicular phase for progesterone and scaled progesterone/estradiol ratio for better interpretation. Same delta direction used accordingly for the BOLD fMRI. All analyses were conducted voxel-wise using a cluster-defining threshold of Z>3.1 (29). For analyses using the two drug cue-reactivity contrasts, cluster-extent thresholds were corrected to p<0.05/2=0.025 for sex and menstrual phase differences separately, and to p<0.05/(2×2)=0.0125 for correlations with cue- and task-induced (post- minus pre- task) drug craving. Analyses pertinent to the reappraisal (and savoring) contrast and correlations with ovarian hormones were more exploratory and therefore corrected to a cluster-extent threshold of p<0.05. In addition to whole-brain analyses, we used an independent anatomical mask (including the entire frontal cortex and striatum) for a restricted search in our regions of interest guided by our previous studies (11, 20) (related results are labeled with “anatomically masked”; see Supplement for additional details of the anatomical mask, neuroimaging data acquisition and preprocessing).

## Results

### Participants

Table 1 illustrates significant sex differences (women>men) in past 30-day use of heroin/cocaine (p=0.005); this effect was not correlated with any of the neuroimaging outcomes of interest and thus was not included as a covariate. No significant sex differences were observed for any of the other drug use or alcohol and smoking variables, demographics, and neuropsychological measures. Group differences were also not observed in any drug use variables between iHUD and iCUD in women. Menstrual phase differences within the subgroup of women underwent two scans were also not observed for dynamic drug use variables (i.e., past 30 days use of drug and days since last drug use).

**Table 1.**
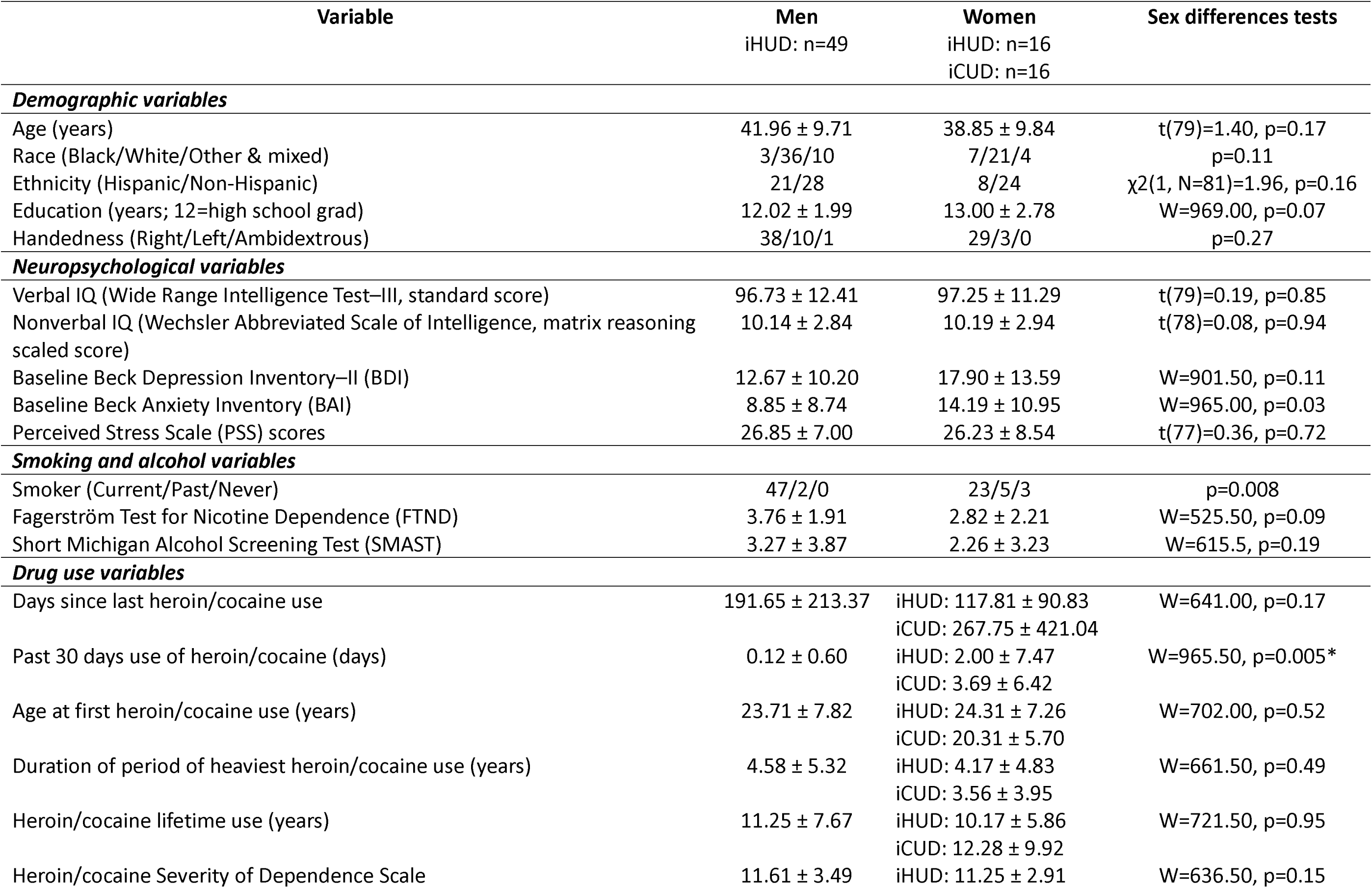
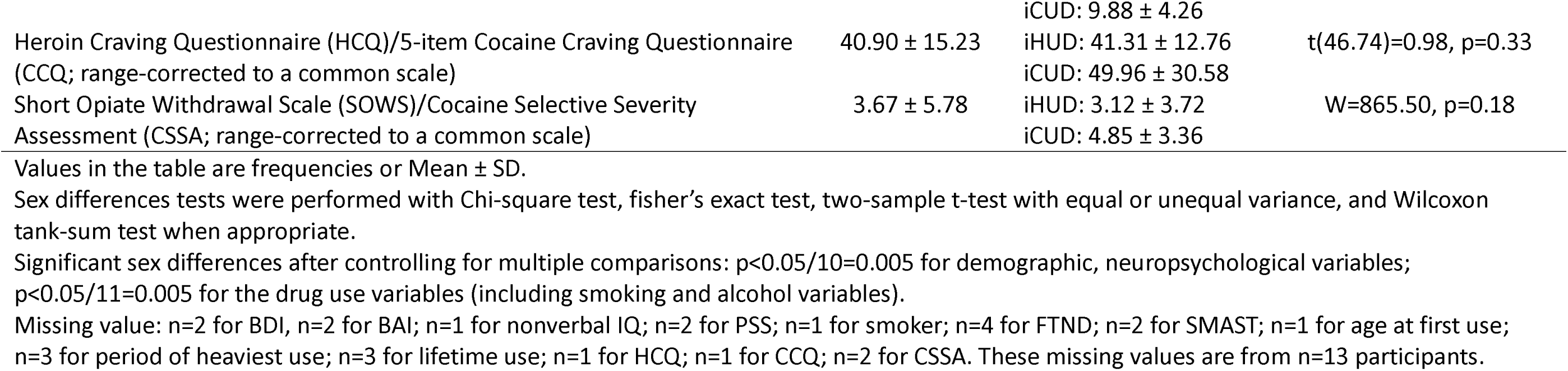
Sample profile.

### Pre- and post-task and post-MRI picture ratings

There were no significant interaction effects with sex or menstrual phase in any of the ratings. See Supplement (Figure S1-S4).

### Sex differences in BOLD fMRI

Whole-brain analyses revealed greater drug cue-reactivity (>look neutral or food) in the medial PFC (mPFC) in women compared to men (Figure 1A). Compared to men, women also showed lower drug reappraisal (>look drug) in the left frontal eye field (FEF)/dlPFC (anatomically masked; Figure 1B). The higher the left dlPFC drug reappraisal, the lower the cue-induced drug craving only in men (anatomically masked; non-significant in women; significant sex differences in slopes via extracted parameter estimates [p=0.042]; Figure 1C).

**Figure 1:**
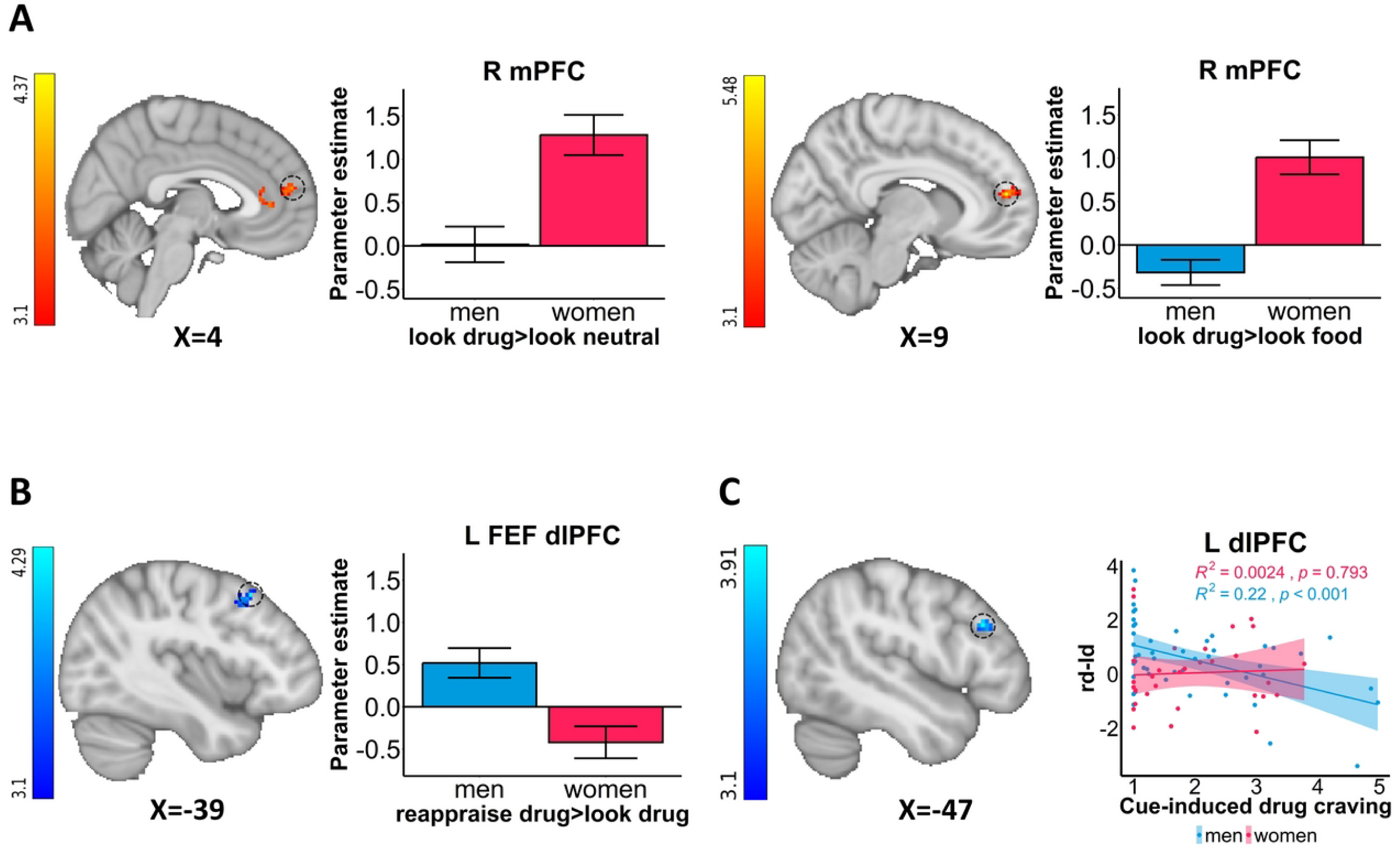
Sex differences in cortico-striatal activity. **A: Sex differences in cortico-striatal drug cue-reactivity.** Women showed higher drug cue-reactivity (>look neutral or look food) in the medial prefrontal cortex (mPFC) than men. **B: Sex differences in cortico-striatal drug reappraisal.** Men showed higher drug reappraisal (>look drug [anatomically masked]) in the frontal eye field (FEF)/dorsolateral PFC (dlPFC) than women**. C: Within-men negative correlation**. Significant negative correlation between the dlPFC (anatomically masked) drug reappraisal and cue-induced drug craving within men only. rd-ld=reappraise drug>look drug; L=left; R=right. For visualization purposes, parameter estimates, depicting blood-oxygen-level-dependent signal, were extracted from corresponding FSL zstat images via 3-mm radius masks centered on Montreal Neurological Institute coordinates from peak activity (black dotted line circles represent the approximate peak coordinates). R^2^ and p values were derived from extracted values. All the error bars denote the mean ± SE.

For completeness, the same analyses were conducted separately within iHUD (men vs. women) and women (iHUD vs. iCUD). Similar sex differences in cortico-striatal activity were observed at a trend level within the smaller sample of iHUD only. There was also no significant group (SUD type within women only) differences that overlapped with these results (see Supplement also for menstrual phase locked sex differences analyses).

### Ovarian hormones

Compared to the late-follicular phase, the mid-luteal phase was characterized by a significantly higher progesterone level (p=0.008), demonstrating the expected progesterone domination (higher scaled progesterone/estradiol ratio: p=0.004). While higher averaged estradiol was observed in the late-follicular phase, there were no significant differences when directly compared to the mid-luteal phase (p=0.56, Figure S5) attributed to the second peak of estradiol level in the menstrual cycle (12, 30). See Supplement for the exploratory hormonal correlations with drug and food cravings.

### Within women menstrual cycle and hormonal effects in BOLD fMRI

Compared to the luteal phase, there was higher FEF/dlPFC drug cue-reactivity (>look food) in the follicular phase (anatomically masked; Figure 2A). These drug cue-reactivity changes between the menstrual phases (Δ) in the IFG and vmPFC (anatomically masked) were positively correlated with Δcue-induced and Δtask-induced drug craving, respectively (Figure 2B & 2C). As compared to the follicular phase, the luteal phase was instead characterized by higher drug reappraisal (>look drug) in the anterior PFC/orbitofrontal cortex (aPFC; Figure 3A) in addition to other areas (Table 2; see Supplement).

**Figure 2:**
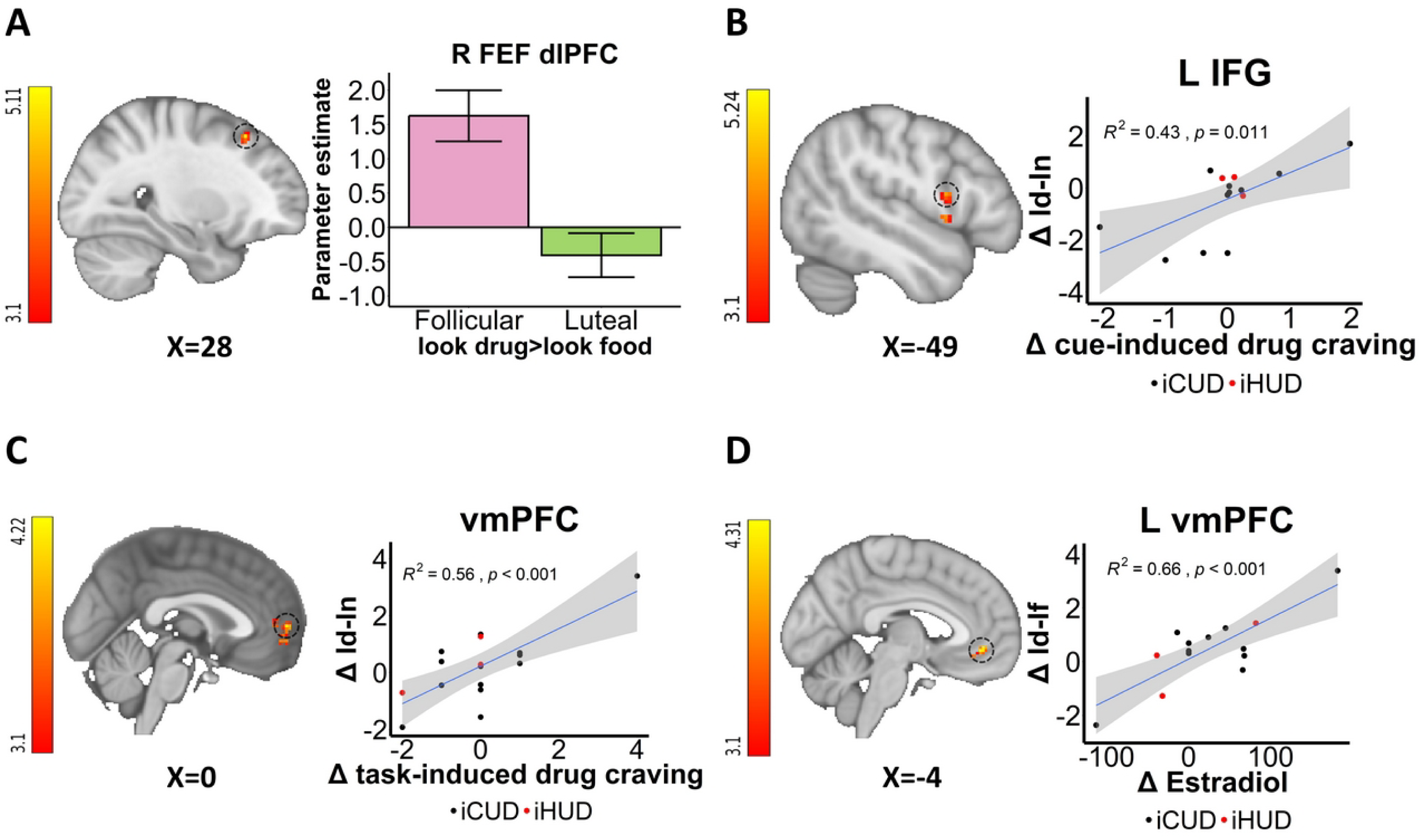
Within women menstrual cycle and hormonal effects of drug cue-reactivity. **A: Menstrual phase differences**. Drug cue-reactivity (>look food [anatomically masked]) in the frontal eye field (FEF)/dorsolateral PFC (dlPFC) is higher during the follicular than the luteal phase. **B-C: Correlations with Δ (between menstrual phase changes) drug craving.** Δdrug cue-reactivity (>look neutral) was positively correlated with Δcue-induced drug craving and Δtask-induced drug craving in inferior frontal gyrus (IFG) and ventromedial prefrontal cortex (vmPFC), respectively. **D: Correlation with Δestradiol.** Δdrug cue-reactivity (>look food [anatomically masked]) was positively correlated with Δestradiol in the vmPFC. iCUD = individuals with cocaine use disorder; iHUD = individuals with heroin use disorder; ld-lf = look drug>look food; ld-ln = look drug>look neutral; L=left; R=right. For visualization purposes, parameter estimates, depicting blood-oxygen-level-dependent signal, were extracted from corresponding FSL zstat images via 3-mm radius masks centered on Montreal Neurological Institute coordinates from peak activity (black dotted line circles represent the approximate peak coordinates). R^2^ and p values were derived from extracted values. All the error bars denote the mean ± SE.

**Figure 3:**
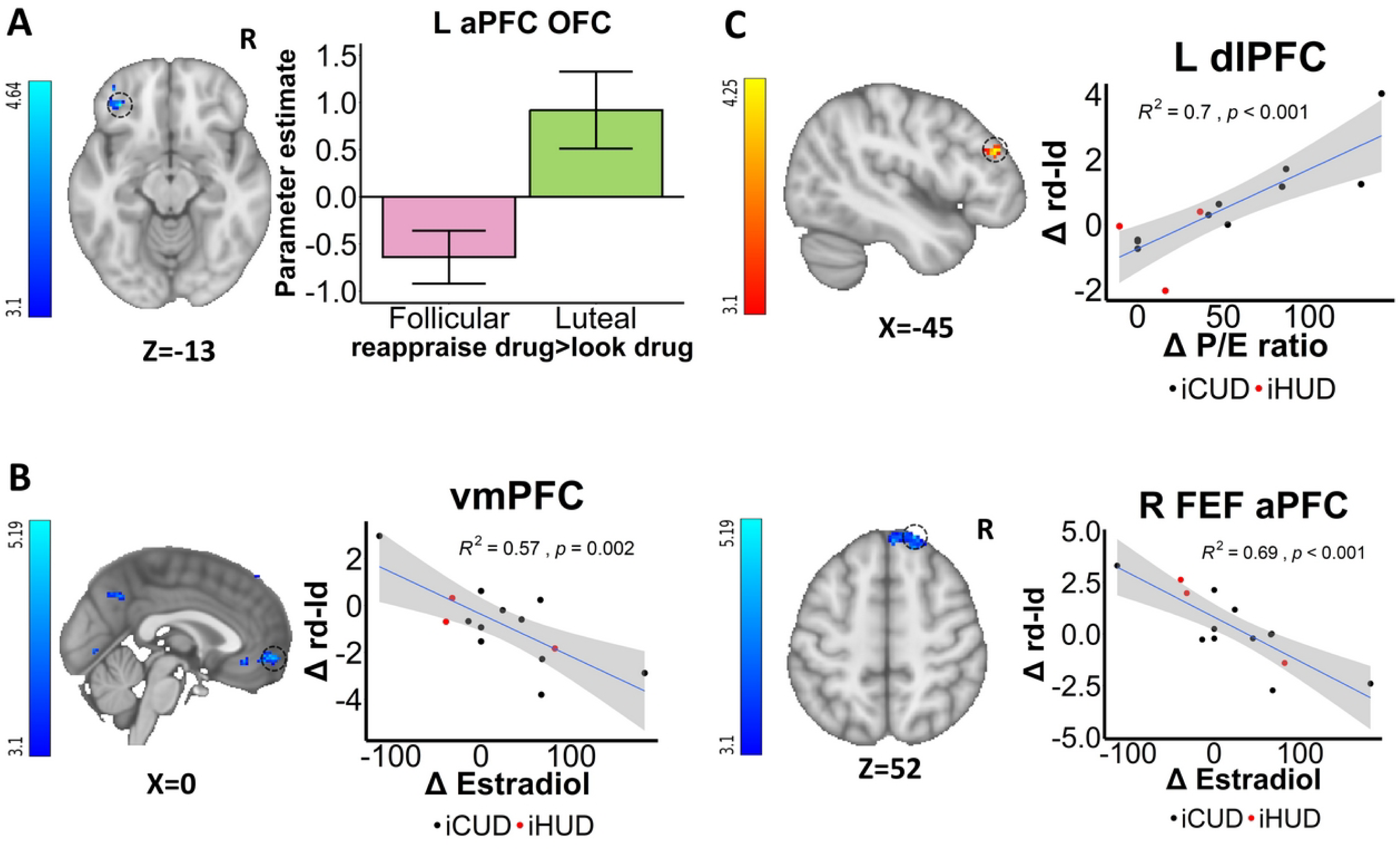
Within women menstrual cycle and hormonal effects of drug reappraisal. **A: Menstrual phase differences**. Drug reappraisal>look drug activity in anterior PFC (aPFC)/orbitofrontal cortex (OFC) was higher during the luteal than the follicular phase. **B: Correlations with Δ (between menstrual phase changes) estradiol.** Δestradiol was negatively correlated with Δdrug reappraisal>look drug activity in the ventromedial prefrontal cortex (vmPFC) and the frontal eye field (FEF)/aPFC. **C: Correlations with Δprogesterone/estradiol (P/E) ratio.** Δdrug reappraisal>look drug activity was positively correlated with ΔP/E ratio in the dorsolateral PFC (dlPFC). iCUD = individuals with cocaine use disorder; iHUD = individuals with heroin use disorder; rd-ld=reappraise drug>look drug; L=left; R=right. For visualization purposes, parameter estimates, depicting blood-oxygen-level-dependent signal, were extracted from corresponding FSL zstat images via 3-mm radius masks centered on Montreal Neurological Institute coordinates from peak activity (black dotted line circles represent the approximate peak coordinates). R^2^ and p values were derived from extracted values. All the error bars denote the mean ± SE.

**Table 2.**
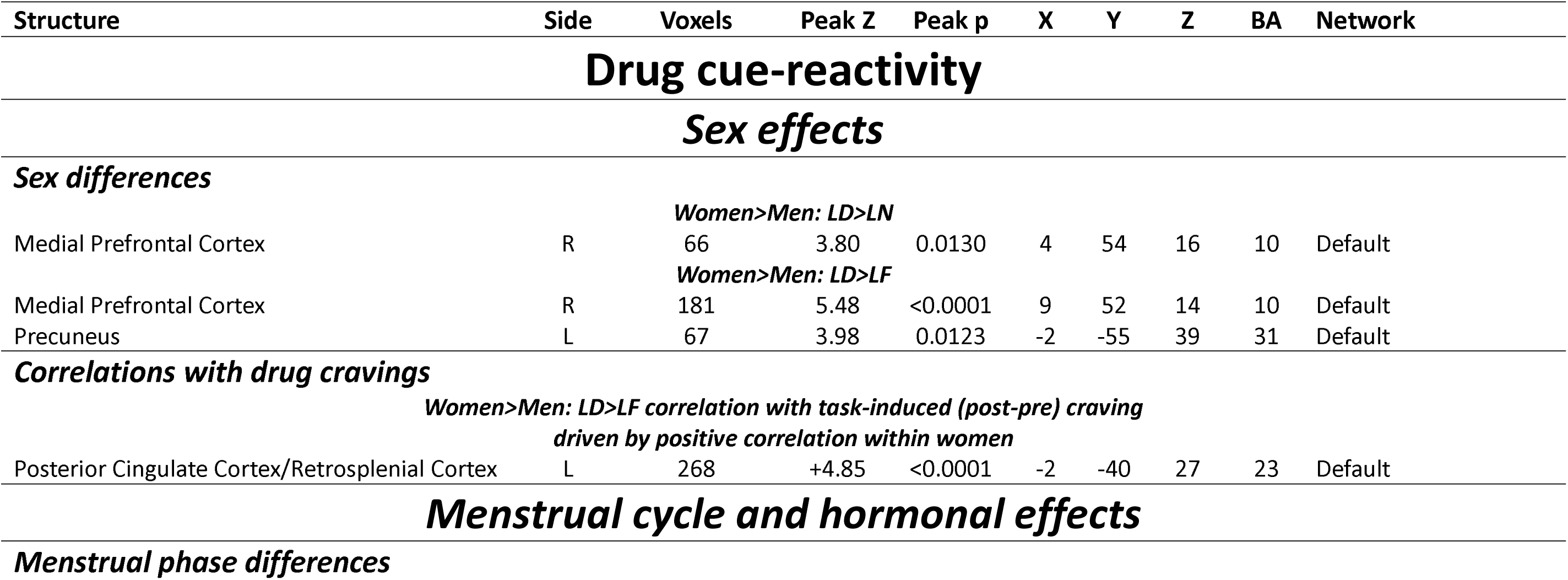
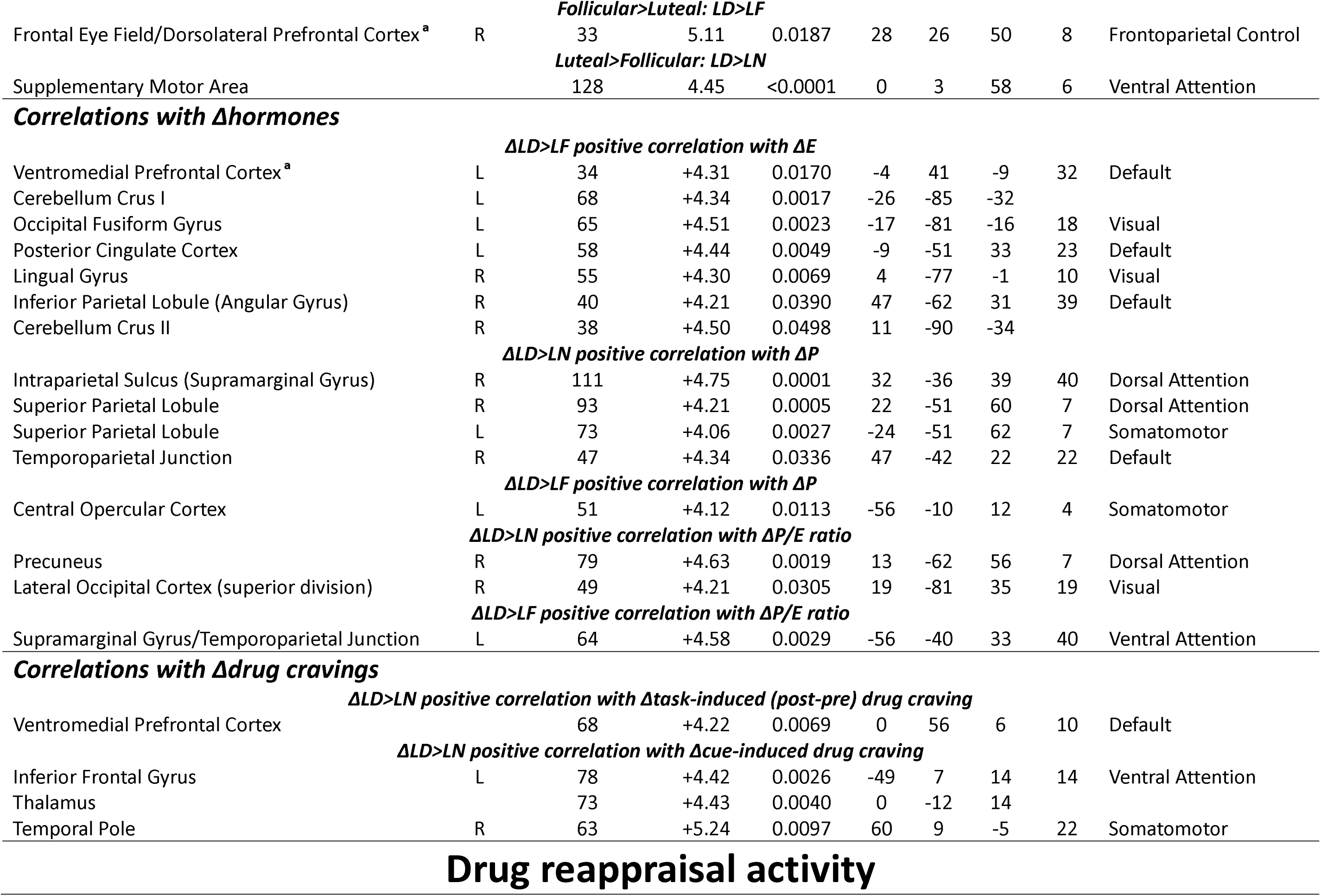
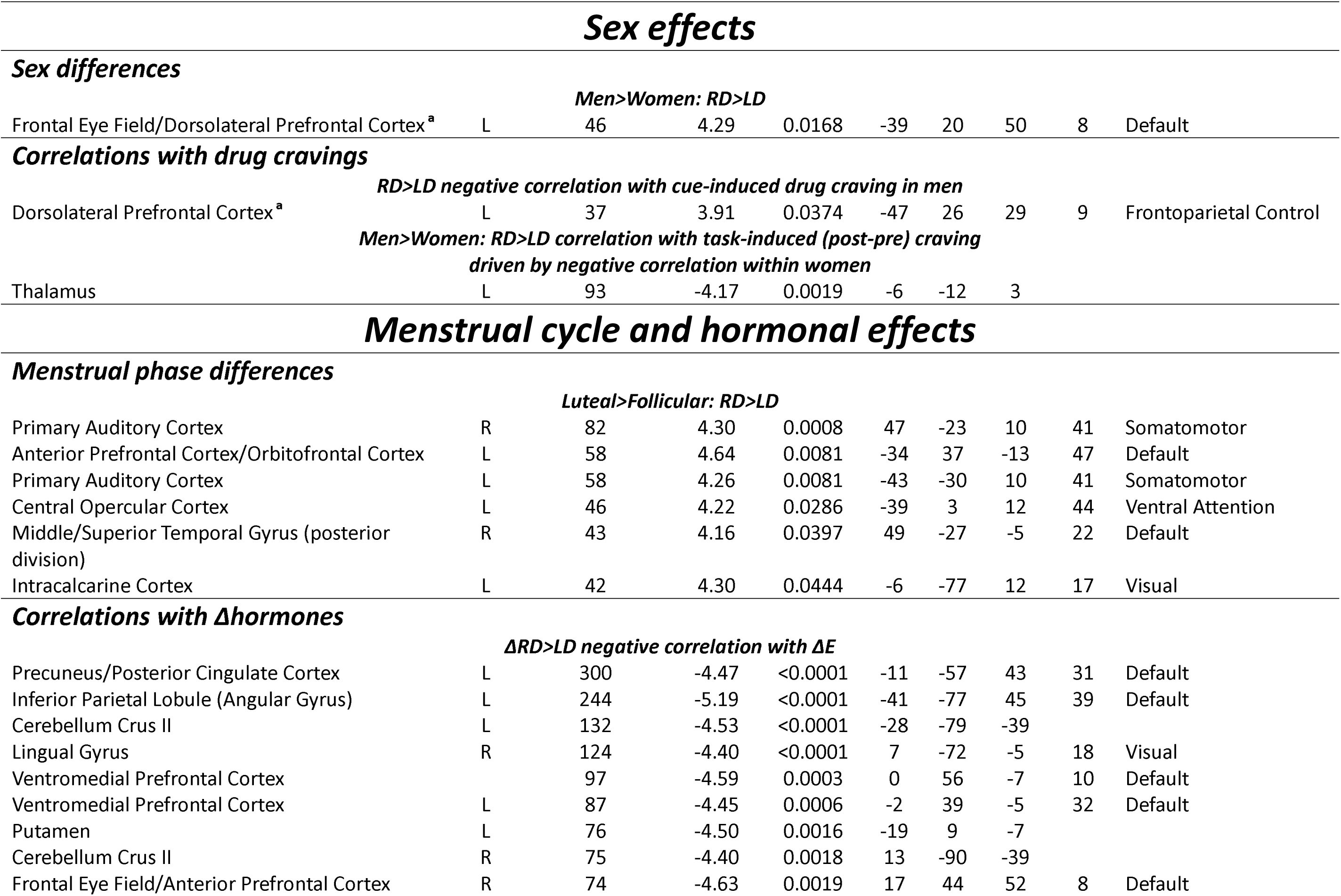
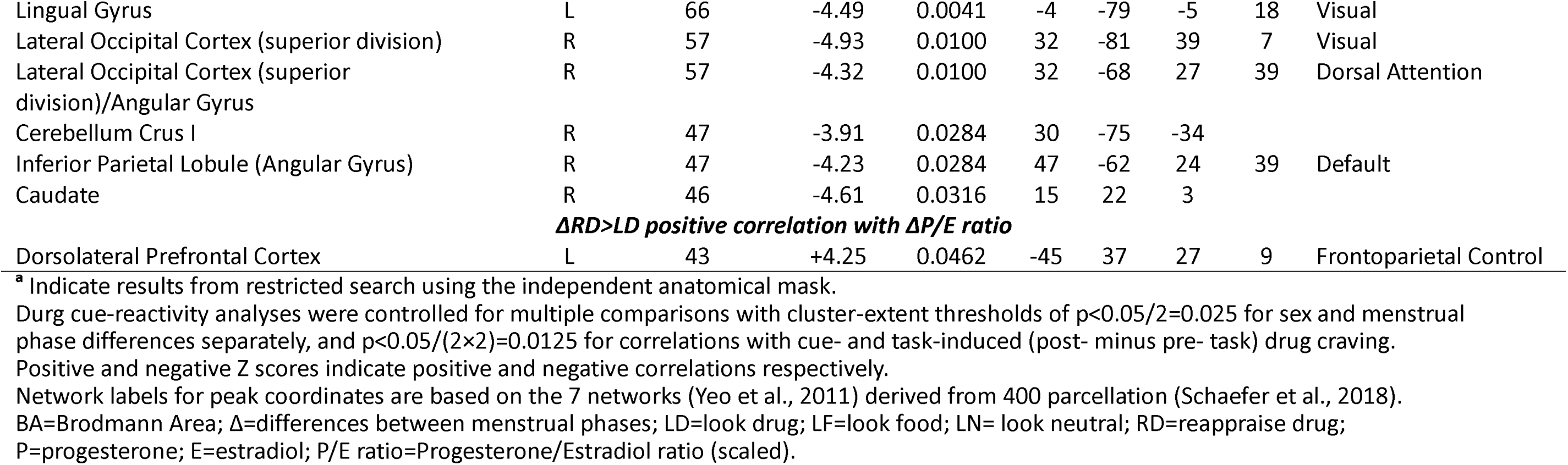
Coordinates for fMRI-BOLD drug cue-reactivity and drug reappraisal results.

Fluctuations in ovarian hormones were directly correlated with these changes in cortico-striatal drug cue-reactivity and drug reappraisal. Specifically, Δestradiol was correlated positively with Δdrug cue-reactivity (>look food) in the vmPFC (anatomically masked; Figure 2D) and negatively with Δdrug reappraisal (>look drug), again, primarily in default mode network regions inclusive of the vmPFC but also in the FEF/aPFC (anatomically masked; Figure 3B) and striatum (including the caudate and putamen; Figure S6B) and other areas (Table 2). In contrast, the Δprogesterone/estradiol ratio was positively correlated with Δdrug reappraisal (>look drug) in the dlPFC (Figure 3C) and other regions (Table 2; see Supplement).

See supplement (Table S1 for savoring contrasts; also Table 2 for contrasts reported in the main text) for results that are outside of our main goals and/or regions of interest (i.e., frontal cortico-striatal areas) for completeness. Also see Figures S7-S9 for summarized (whole-brain and anatomically masked) cortical BOLD fMRI activation for menstrual cycle and hormonal effects.

## Discussion

We first demonstrated elevated mPFC drug cue-reactivity in women and higher FEF/dlPFC drug reappraisal as correlated with lower cue-induced drug craving in men. Additionally, significant menstrual cycle effects were found, where drug cue-reactivity was higher in the FEF/dlPFC during the follicular phase and drug reappraisal was higher in the aPFC/orbitofrontal cortex during the luteal phase. Within-subject correlations on changes between menstrual phases showed that ΔvmPFC drug cue-reactivity correlated with both higher Δestradiol and Δtask- induced drug craving; a similar correlation was observed for the IFG. Conversely, Δdrug reappraisal correlated with lower Δestradiol (in the vmPFC, FEF/aPFC, and striatum) and higher Δprogesterone/estradiol ratio (in the dlPFC).

Consistent with findings in people with problem alcohol use (13, 14) but opposite to a study in iCUD (31, 32) (potentially due to different task stimuli used [imaginary scripts vs. pictures] and the regulation conditions in our task), we found higher mPFC drug cue-reactivity in women with HUD or CUD compared to men with HUD. In the men, we observed higher drug reappraisal in the FEF/dlPFC, consistent with studies in healthy individuals (21, 22), further linking this activity to lower cue-induced drug craving in our HUD men. This result is aligns with the general role of the dlPFC in top-down cognitive control (e.g., during emotion regulation (33)) and its specific role in craving regulation (e.g., in current smokers (34)). The absence of a similar correlation in our SUD women further suggests a potentially sexually dimorphic mechanism, where men may better mobilize top-down control resources during cognitive reappraisal to reduce drug cue-reactivity and the associated experience of craving, while in women absence of these effects may suggest more susceptibility/vulnerability specifically during drug cue exposure.

These effects may be influenced hormonally. Indeed, enhanced drug cue-reactivity was observed during the follicular phase (FEF/dlPFC) as associated with increased Δdrug craving (IFG and vmPFC) and Δestradiol (vmPFC). The luteal phase instead showed enhanced drug reappraisal (aPFC) as associated with reduced Δestradiol (FEF/aPFC and vmPFC and striatum [encompassing putamen and caudate], potentially attributed to estrogen’s positive modulation effects on the dopamine system, specific to females (35)) and increased Δprogesterone/estradiol ratio (dlPFC). These results highlight a unique role for the vmPFC (menstrual phase differences in both drug cue-reactivity and reappraisal associated in the opposite, and expected, directions with respective estradiol changes and with craving changes for the former), suggesting this default mode network region may be a menstrual/hormone-modulated neural marker for addiction vulnerability/risk vs. resilience in women. Therefore, the vmPFC’s role in subjective valuation (36) and the generation of affective meaning (37) during drug cue exposure and its regulation could be targeted (for its attenuation in the former and enhancement in the latter) in future intervention efforts. These results also highlight direct hormonal effects where, in contrast to estrogen, progesterone appears to have a protective effect (38, 39). In our study this conclusion is supported by the higher drug reappraisal during the luteal phase and by the positive association between the Δprogesterone/estradiol ratio with Δdrug reappraisal in executive and cognitive control regions (40) (e.g., the dlPFC, a region within the frontoparietal control network). These anterior lateral PFC regions’ role in emotion regulation (33, 41) could therefore be targeted for enhancement (e.g., with transcranial direct current stimulation (42)). Abnormalities within and between the default mode network and attention and executive control networks have also been associated with facilitating craving and relapse in SUD (43), and hence the dynamic co-activation of these networks could be a further treatment target.

Some limitations need to be acknowledged and future directions discussed: 1) The current study lacked a group of men with CUD, preventing a direct comparison between substances. However, using an fMRI task we adapted to be relevant to both iCUD and iHUD, our results suggest some generalizability across substances (note also stability of results when excluding participants who are comorbid for both HUD and CUD, see Supplement). Nevertheless, future efforts should fill this gap (also need more samples for CUD vs. HUD effect within women due to comorbidity on these substances; see Supplement); 2) While we uncovered significant menstrual cycle/hormonal effects with a small sample size, it is important to replicate these findings with a larger sample size and more balanced groups. Adding a group of healthy control women (and/or a second scan in men) for reference of the hormonal effects would also increase the validity of the current findings; 3) Our study targeted the mid-follicular and mid-luteal phases (when estradiol is relatively high, Figure S5), potentially contributing to some negative results. Future studies should therefore assess the additional early-follicular phase (low estradiol and progesterone), especially as the strongest ventral striatal drug cue-reactivity was observed when comparing this phase to the late-follicular and mid-luteal phases in nicotine-dependent women (12) ; 4) Stress-induced drug cue-reactivity and fluctuations in the stress and reward brain circuits’ function during the menstrual cycle were reported in women with CUD, linked to fluctuations in cortisol in addition to ovarian hormones (31, 44). Therefore, measuring the cortisol hormone could be informative for future research; 5) Long-term exposure to opioids or opioid replacement therapy (e.g., methadone and buprenorphine) could lead to hypogonadism (e.g., deficiencies in estradiol and progesterone) (45). Indeed, some of our participants with urine positive for methadone or buprenorphine showed very low levels (below sensitivity level) of progesterone or estradiol, suggesting that this factor is important for future study. We could also not equate the women subgroups on this factor, although we note lack of differences between the CUD and HUD women; and 6) While no participants in our analyses reported current or past gynecological problems that require medical attention nor recent alterations in menstrual cycle patterns, future studies will benefit from a more extensive medical examination to rule out potential conditions (e.g., endometriosis or polycystic ovary syndrome) that could alter hormonal profiles.

To the best of our knowledge, this is the first clinical neuroimaging study simultaneously examining sex differences, menstrual phase, and hormonal effects in cortico-striatal drug cue-reactivity and its regulation in SUD. Although the sex differences reported here remain to be reconciled with the higher overdose mortality rates in men at the epidemiological level (6), our findings offer valuable and timely insights into the hormone-modulated vulnerability and resilience factors in women with SUD. Specifically, our results could inform translational efforts to develop precisely timed, hormonally informed treatments. For example, we recommend targeting the follicular phase to reduce drug cue-reactivity by enhancing the executive control functions (e.g., cognitive reappraisal and/or savoring of an alternative reward, see Supplement), which may reduce craving and ultimately improve treatment outcomes in women with SUD.

## Data Availability

The data that support the findings of this study are available from the corresponding author, upon reasonable request.

## References

1. Becker JB, McClellan ML, Reed BG: Sex differences, gender and addiction. Journal of Neuroscience Research 2017; 95:136–147

2. Towers EB, Williams IL, Qillawala EI, et al.: Sex/Gender Differences in the Time-Course for the Development of Substance Use Disorder: A Focus on the Telescoping Effect. Pharmacol Rev 2023; 75:217–249

3. Lynch WJ, Roth ME, Carroll ME: Biological basis of sex differences in drug abuse: preclinical and clinical studies. Psychopharmacology 2002; 164:121–137

4. Butelman ER, Chen CY, Brown KG, et al.: Escalation of drug use in persons dually diagnosed with opioid and cocaine dependence: Gender comparison and dimensional predictors. Drug Alcohol Depend 2019; 205:107657

5. Nicolas C, Zlebnik NE, Farokhnia M, et al.: Sex Differences in Opioid and Psychostimulant Craving and Relapse: A Critical Review. Pharmacol Rev 2022; 74:119–140

6. Butelman ER, Huang Y, Epstein DH, et al.: Overdose mortality rates for opioids and stimulant drugs are substantially higher in men than in women: state-level analysis. Neuropsychopharmacol 2023; 48:1639–1647

7. Sofuoglu M, Dudish-Poulsen S, Nelson D, et al.: Sex and menstrual cycle differences in the subjective effects from smoked cocaine in humans. Experimental and Clinical Psychopharmacology 1999; 7:274–283

8. Evans SM, Haney M, Foltin RW: The effects of smoked cocaine during the follicular and luteal phases of the menstrual cycle in women. Psychopharmacology 2002; 159:397–406

9. Baker NL, Gray KM, Ramakrishnan V, et al.: Increases in endogenous progesterone attenuate smoking in a cohort of nontreatment seeking women: An exploratory prospective study. Addiction Biology 2021; 26:e12918

10. Moningka H, Lichenstein S, Worhunsky PD, et al.: Can neuroimaging help combat the opioid epidemic? A systematic review of clinical and pharmacological challenge fMRI studies with recommendations for future research. Neuropsychopharmacol 2019; 44:259–273

11. Zilverstand A, Huang AS, Alia-Klein N, et al.: Neuroimaging Impaired Response Inhibition and Salience Attribution in Human Drug Addiction: A Systematic Review. Neuron 2018; 98:886–903

12. Franklin TR, Spilka NH, Keyser H, et al.: Impact of the natural hormonal milieu on ventral striatal responses to appetitive cigarette smoking cues: A prospective longitudinal study. Drug and Alcohol Dependence Reports 2022; 5:100119

13. Maxwell AM, Brucar LR, Zilverstand A: A Systematic Review of Sex/Gender Differences in the Multi- dimensional Neurobiological Mechanisms in Addiction and Their Relevance to Impulsivity [Internet]. Curr Addict Rep 2023; [cited 2023 Dec 6] Available from: 10.1007/s40429-023-00529-9

14. Claus ED, Blaine SK, Witkiewitz K, et al.: Sex moderates effects of alcohol and cannabis co-use on alcohol and stress reactivity. Alcoholism: Clinical and Experimental Research 2022; 46:530–541

15. Radoman M, Fogelman N, Lacadie C, et al.: Neural Correlates of Stress and Alcohol Cue-Induced Alcohol Craving and of Future Heavy Drinking: Evidence of Sex Differences. AJP 2024; 181:412–422

16. Li Q, Li W, Wang H, et al.: Predicting subsequent relapse by drug-related cue-induced brain activation in heroin addiction: an event-related functional magnetic resonance imaging study. Addiction Biology 2015; 20:968–978

17. Zilverstand A, Parvaz MA, Moeller SJ, et al.: Cognitive interventions for addiction medicine: Understanding the underlying neurobiological mechanisms. Prog Brain Res 2016; 224:285–304

18. Bryan MA, Mallik D, Cochran G, et al.: Mindfulness and Savoring: A Commentary on Savoring Strategies and Their Implications for Addiction Treatment. Substance Use & Misuse 2022; 57:822– 826

19. Garland EL, Atchley RM, Hanley AW, et al.: Mindfulness-Oriented Recovery Enhancement remediates hedonic dysregulation in opioid users: Neural and affective evidence of target engagement. Sci Adv 2019; 5:eaax1569

20. Huang Y, Ceceli AO, Kronberg G, et al.: Association of Cortico-Striatal Engagement During Cue Reactivity, Reappraisal, and Savoring of Drug and Non-Drug Stimuli With Craving in Heroin Addiction. AJP 2024; 181:153–165

21. Mak AKY, Hu Z, Zhang JXX, et al.: Sex-related differences in neural activity during emotion regulation. Neuropsychologia 2009; 47:2900–2908

22. Domes G, Schulze L, Böttger M, et al.: The neural correlates of sex differences in emotional reactivity and emotion regulation. Hum Brain Mapp 2010; 31:758–769

23. Albert KM, Boyd BD, Taylor WD, et al.: Differential effects of estradiol on neural and emotional stress response in postmenopausal women with remitted Major Depressive Disorder. Journal of Affective Disorders 2021; 293:355–362

24. Rehbein E, Kogler L, Hornung J, et al.: Estradiol administration modulates neural emotion regulation. Psychoneuroendocrinology 2021; 134:105425

25. Ahumada-Méndez F, Lucero B, Avenanti A, et al.: Affective modulation of cognitive control: A systematic review of EEG studies. Physiology & Behavior 2022; 249:113743

26. Zelionkaitė I, Gaižauskaitė R, Uusberg H, et al.: The levonorgestrel-releasing intrauterine device is related to early emotional reactivity: An ERP study. Psychoneuroendocrinology 2024; 162:106954

27. Woolrich MW, Ripley BD, Brady M, et al.: Temporal Autocorrelation in Univariate Linear Modeling of FMRI Data. NeuroImage 2001; 14:1370–1386

28. Beckmann CF, Jenkinson M, Smith SM: General multilevel linear modeling for group analysis in FMRI. NeuroImage 2003; 20:1052–1063

29. Eklund A, Nichols TE, Knutsson H: Cluster failure: Why fMRI inferences for spatial extent have inflated false-positive rates. Proc Natl Acad Sci U S A 2016; 113:7900–7905

30. Becker JB, Arnold AP, Berkley KJ, et al.: Strategies and Methods for Research on Sex Differences in Brain and Behavior. Endocrinology 2005; 146:1650–1673

31. Potenza MN, Hong KA, Lacadie CM, et al.: Neural Correlates of Stress-Induced and Cue-Induced Drug Craving: Influences of Sex and Cocaine Dependence. AJP 2012; 169:406–414

32. Smith K, Lacadie CM, Milivojevic V, et al.: Sex differences in neural responses to stress and drug cues predicts future drug use in individuals with substance use disorder. Drug and Alcohol Dependence 2023; 244:109794

33. Buhle JT, Silvers JA, Wager TD, et al.: Cognitive Reappraisal of Emotion: A Meta-Analysis of Human Neuroimaging Studies. Cerebral Cortex 2014; 24:2981–2990

34. Kober H, Mende-Siedlecki P, Kross EF, et al.: Prefrontal–striatal pathway underlies cognitive regulation of craving. PNAS 2010; 107:14811–14816

35. Becker JB: Direct effect of 17β-estradiol on striatum: Sex differences in dopamine release. Synapse 1990; 5:157–164

36. Levy I, Snell J, Nelson AJ, et al.: Neural Representation of Subjective Value Under Risk and Ambiguity. Journal of Neurophysiology 2010; 103:1036–1047

37. Roy M, Shohamy D, Wager TD: Ventromedial prefrontal-subcortical systems and the generation of affective meaning. Trends in Cognitive Sciences 2012; 16:147–156

38. Becker JB, Hu M: Sex differences in drug abuse. Frontiers in Neuroendocrinology 2008; 29:36–47

39. Lynch WJ: Acquisition and maintenance of cocaine self-administration in adolescent rats: effects of sex and gonadal hormones. Psychopharmacology 2008; 197:237–246

40. Menon V, D’Esposito M: The role of PFC networks in cognitive control and executive function. Neuropsychopharmacol 2022; 47:90–103

41. Bo K, Kraynak TE, Kwon M, et al.: A systems identification approach using Bayes factors to deconstruct the brain bases of emotion regulation. Nat Neurosci 2024; 27:1–13

42. Gaudreault P-O, Sharma A, Datta A, et al.: A double-blind sham-controlled phase 1 clinical trial of tDCS of the dorsolateral prefrontal cortex in cocaine inpatients: Craving, sleepiness, and contemplation to change. European Journal of Neuroscience 2021; 53:3212–3230

43. Zhang R, Volkow ND: Brain default-mode network dysfunction in addiction. NeuroImage 2019; 200:313–331

44. Fox HC, Hong KA, Paliwal P, et al.: Altered levels of sex and stress steroid hormones assessed daily over a 28-day cycle in early abstinent cocaine-dependent females. Psychopharmacology 2008; 195:527–536

45. Antony T, Alzaharani SY, El-Ghaiesh SH: Opioid-induced hypogonadism: Pathophysiology, clinical and therapeutics review. Clinical and Experimental Pharmacology and Physiology 2020; 47:741– 750

