## supplementary material for "Sex and hormonal effects on drug cue-reactivity and its regulation in human addiction"

**Exclusion criteria**

Exclusion criteria for all participants were the following: 1) DSM-5 diagnosis for schizophrenia or developmental disorder; 2) Head trauma with loss of consciousness (>30 min); 3) History of neurological disease of central origin; 4) Cardiovascular, metabolic, endocrinological, oncological, autoimmune, and current infectious diseases including Hepatitis B and C or HIV/AIDS; 5) Metal implants or other magnetic resonance imaging (MRI) contraindications (including pregnancy). We did not exclude for DSM-5 diagnosis of a drug use disorder other than opiates/stimulants as long as heroin/cocaine, respectively, was the primary drug of choice/reason for treatment-seeking since individuals with heroin/cocaine use disorder (HUD; CUD) commonly use alcohol, amphetamines, benzodiazepines, other sedatives, and marijuana in addition to heroin/cocaine.

**Participants**

All but one woman iHUD and nine women iCUD were treatment-seeking individuals. They were recruited from medication-assisted inpatient or outpatient addiction treatment facilities in New York City and had been medically (methadone or buprenorphine) stabilized for at least 2 weeks. Non-treatment-seeking individuals were recruited from the same area.

Clinical diagnostic interview encompassed the Mini-International Neuropsychiatric Interview(1) for Diagnostic and Statistical Manual of Mental Disorder (DSM)-5 criteria and the Addiction Severity Index,(2) a semi-structured instrument that captures drug use history and severity. Severity of drug dependence, craving, and withdrawal symptoms were assessed with the Severity of Dependence Scale(3) for all participants, the Heroin Craving Questionnaire (a modified version of the Cocaine Craving Questionnaire) for iHUD(4, 5) and the 5-item cocaine craving questionnaire (range-corrected to a common scale for comparisons) for iCUD(4), and the Short Opiate Withdrawal Scale(6) for iHUD, and Cocaine Selective Severity Assessment (range-corrected to a common scale for comparisons)(7) for iCUD, respectively. A brief physical examination, including heart rate, blood pressure, urine drug toxicology, alcohol concentration (via saliva testing strips), and breath carbon monoxide levels, and a review of medical history were also performed by trained research staff. All women were tested for pregnancy via urine testing strips at each MRI day.

All 16 women (iCUD=13, iHUD=3) who underwent two MRI scans to examine menstrual cycle effects were screened with a comprehensive menstrual history questionnaire. This assessment included their current menstrual cycle pattern (e.g., average cycle length over the past 3 months), current or past 6 months contraceptive use, menstrual symptoms, and past menstrual history.

Other comorbidities for the 49 men and 16 women with HUD were depression (N=32), post-traumatic stress disorder (N=12), general anxiety (N=4), alcohol use disorder (N=16), other than crack/cocaine stimulants use disorder (N=2), cannabis use disorder (N=4), and tranquilizer use disorder (N=11). In the 49 men with HUD, 17 also had CUD (7 in early remission, 3 in sustained remission, and 7 with current CUD). In the 16 women with HUD, 3 also had CUD (1 in early remission and 2 current CUD).

For the 16 women with CUD, other comorbidities were depression (N=7), post-traumatic stress disorder (N=3), alcohol use disorder (N=4), other than crack/cocaine stimulant use disorder (N=3), hallucinogens use disorder (N=1), and cannabis use disorder (N=2). In these 16 women with HUD, 5 also had opiate use disorder (OUD; 1 in early remission, 1 in sustained remission, and 3 with current OUD).

Primary routes of administration for the 49 men and 16 women with HUD were intravenous injection (N=39), non-intravenous injection (N=1), intranasal (N=21), smoking/inhaling (N=3), and oral (N=1). For the 16 women with CUD these were intravenous injection (N=3), intranasal (N=11), and smoking/inhaling (N=2). This pattern of results was similar for the subjects comorbid for HUD/OUD and CUD. Across all the participants, urine toxicology assessment on the first MRI scan day (for sex differences effect) indicated the presence of methadone (N=59), buprenorphine (N=8), methadone and buprenorphine (N=1; participant switched from suboxone to methadone 4 days before the scan), benzodiazepines (N=1), cocaine (N=6), opiates (N=2), morphine (N=1), antidepressants (N=19), methamphetamine (N=1), amphetamine (N=2), and marijuana/THC (N=4). One datapoint for urine toxicology data was missing. Six iHUD and six iCUD had used heroin and cocaine, respectively, during the 30 days preceding the scan.

For the 16 women (iCUD=13, iHUD=3) who were studied for the menstrual cycle effect, urine toxicology assessment on the second MRI scan day indicated the presence of methadone (N=6), buprenorphine (N=1), antidepressants (N=5), amphetamine (N=2), morphine (N=1), and marijuana/THC (N=1). One datapoint for urine toxicology data was missing. One iHUD and five iCUD had used heroin and cocaine, respectively, during the 30 days preceding the scan.

**fMRI Task instructions**

Task instructions for the look, reappraise, and savor conditions were adapted from previous studies(8, 9). During the look condition, participants were instructed to “keep viewing the picture normally”. During the reappraise condition, participants were instructed to reduce their emotional reactivity to the heroin/cocaine pictures in three practice trials, each providing a different strategy: 1) “Try to imagine that the scenario is not real, that it is from a movie, and these are all actors”; 2) “Try to imagine that the heroin/cocaine is not real, that it is just a prop”; 3) “You can focus on how this is just a picture, and tell yourself that it is not real heroin/cocaine”. During the savor condition, participants were instructed to increase their emotional reactivity to the food pictures in three other practice trials, each providing a different savoring strategy: 1) “You can imagine that you are holding the food in the picture, and feeling the weight of it in your hands, enjoying its pleasant smell”; 2) “You can focus on how good the food looks or imagine how good it would taste, savoring the delicious taste of the food”; 3) “Imagine the sensation of how it would feel in your mouth or how it feels once you’ve eaten it”. Participants were instructed to verbally describe their reappraisal and savoring strategies during these practice trials to ensure task comprehension, but they were instructed to refrain from speaking during the fMRI task trials in the scanner.

**Pre- and post-task ratings**

Participants provided drug (heroin or cocaine) and food craving ratings, as well as a ratings of motivation to complete the task on a 10-point scale immediately before the functional MRI (fMRI) cue-reactivity task (i.e., “Please rate how strong your desire for heroin [or cocaine] is currently on a scale of 0-9”, “Please rate how strong your desire for food is currently on a scale of 0-9”, and “Please rate your motivation to complete this task on a scale of 0-9”). Immediately after the cue-reactivity task, in addition to the same three questions, participants were asked to provide self-evaluation of the difficulty and effectiveness of their reappraisal and savoring performance on a 10-point scale (i.e., “How difficult did you find it to decrease your emotional reactivity to the heroin [or cocaine] pictures during this task?”, “How well do you think you decreased your emotional reactivity to the heroin [or cocaine] pictures during this task?”, “How difficult did you find it to increase your emotional reactivity to the food pictures during this task?” and “How well do you think you increased your emotional reactivity to the food pictures during this task?”).

*Sex differences analyses*

To analyze pre- and post-task drug/food craving ratings between men and women, a 2 (sex: men/women) by 2 (time: pre/post) by 2 (image: drug/food) mixed analysis of variance (ANOVA) was conducted. We found significant main effects of time [post>pre: F(1,77) = 11.43, p = 0.001] and image [food>drug: F(1,77) = 54.42, p < 0.001], indicative of higher craving ratings after the task and higher food than drug craving across all subjects. No significant main effect of sex or interaction effects were found (p>0.553). A 2 (sex: men/women) by 2 (time: pre/post) mixed ANOVA for motivation ratings showed no significant main or interaction effects (p>0.204). Regarding self-evaluated emotion regulation performance, four two sample t-tests for difficulty and effectiveness were conducted for reappraisal and savoring separately to test for sex differences. No sex differences were found in the emotion regulation performance (p>0.169). Overall, these results suggest that there were no sex differences in craving ratings, task motivation or self-evaluated difficulty and effectiveness in reappraisal and savoring (Figure S1).

*Menstrual cycle analyses*

To analyze pre- and post-task drug/food craving ratings between menstrual phases, a 2 (menstrual phases: follicular/luteal) by 2 (time: pre/post) by 2 (image: drug/food) within-subject analysis of variance (ANOVA) was conducted. Similarly to the above results, we found significant main effects of time [post>pre: F(1,15) = 8.84, p = 0.009] and image [food>drug: F(1,15) = 10.57, p = 0.005]. No significant interaction effects or main effect of menstrual phase were found (p>0.247). A 2 (menstrual phases: follicular/luteal) by 2 (time: pre/post) within-subject ANOVA for motivation ratings showed no significant main or interaction effects (p>0.437). Regarding self-evaluated emotion regulation performance, four paired t-tests for difficulty and effectiveness were conducted for reappraisal and savoring separately to test for menstrual phase differences. No significant menstrual phase differences were found in the emotion regulation performance (p>0.688). Overall, these results suggest that there were no menstrual phase differences in craving ratings, task motivation or self-evaluated difficulty and effectiveness in reappraisal and savoring (Figure S2).

**Post-MRI picture ratings**

After the MRI, participants provided image-specific valence and arousal ratings on the drug, food, and neutral images, as well as cue-induced craving (i.e., wanting) ratings on the drug and food images on a subset of images presented in the task. The images to be rated were pseudorandomized based on the odd or even number combination of the participant ID and session number. For valence ratings, participants were asked ‘How pleasant do you find the above picture?’ on a 5-point scale from 1 (very unpleasant) to 5 (very pleasant). For arousal, participants were asked “How emotional do you feel about the above picture?” on a 5-point scale from 1 (calm, no emotion) to 5 (extremely emotional). For wanting, participants were asked “How strong is your desire to use the above substance (or food)?” on a 5-point scale from 1 (no desire) to 5 (extreme desire).

*Sex differences analyses*

To test for potential sex differences between valence and arousal ratings for food, drug and neutral images, two separate 2 (sex: men/women) by 3 (images: drug/food/neutral) mixed ANOVAs were conducted. For valence, there was a significant main effect of image [food>neutral>drug: F(1.32,103.06) = 128.66, p < 0.001] and a trend for main effect of sex [women>men: F(1,78) = 3.92, p=0.051]. There was no significant interaction (p = 0.137). For arousal, the main effects of image [food=drug>neutral: F(1.34,104.75) = 16.25, p < 0.001] and sex [men>women: F(1,78) = 5.54, p = 0.021] were significant. The interaction effect between sex and image was not significant (p = 0.079). A 2 (sex: men/women) by 2 (image: drug/food) mixed ANOVA on the wanting/cue-induced craving ratings showed a significant main effect for image [food>drug: F(1,78) = 32.21, p < 0.001] with no significant main effect of sex (p = 0.627) or a sex × image interaction (p = 0.657) (Figure S3). These results indicate that the food cues were more positively valanced and elicited stronger craving responses than the drug cues, although they evoked the same arousal across participants. Men provided higher arousal ratings overall, while there was a trend for higher valence ratings in women. However, the interaction effects with sex were not significant for both arousal and valence across image types, and hence these ratings were not used as covariates in the analyses reported in the main text. There were no other effects in these ratings.

*Menstrual cycle analyses*

To test potential menstrual phase differences between the valence and arousal ratings for food, drug and neutral images, two separate 2 (menstrual phases: follicular/luteal) by 3 (images: drug/food/neutral) within-subject ANOVAs were conducted. Similarly to the above results, for valence, there was a significant main effect of image [food>neutral>drug: F(1.14,14.79) = 16.52, p < 0.001]. There was no significant main effect of menstrual phase or interaction effect between menstrual phase and image (p>0.169). For arousal, the main effect of image [drug>food=neutral: F(1.87,24.27) = 10.83, p < 0.001] was significant while the main effect of menstrual phase or interaction effect between menstrual phase and image were not significant (p>0.390). A 2 (menstrual phases: follicular/luteal) by 2 (image: drug/food) within-subject ANOVA on the wanting/cue-induced craving ratings showed a significant main effect for image [food>drug: F(1,13) = 5.61, p = 0.034] with no significant main effect of menstrual phase (p = 0.394) or a menstrual cycle × image interaction (p = 0.302) (Figure S4). These results indicate no significant main or interaction menstrual cycle effects for picture ratings of valence, arousal and craving.

**MRI data acquisition and preprocessing**

The MRI protocol was optimized to be Human Connectome Project compatible(10) and the data was collected on a Siemens 3-T Skyra scanner (Siemens Healthcare, Erlangen, Germany) using a 32-channel head coil. Anatomical T1-weighted structural images were acquired using the following parameters: 3D MPRAGE (Magnetization-Prepared Rapid Gradient-Echo) sequence with FOV of 256 × 256 × 179 mm3, 0.8 mm isotropic resolution, TR/TE/TI = 2400/2.07/1000 ms, 8° flip angle with binomial (1, −1) fat saturation, 240 Hz/pixel bandwidth, 7.6 ms echo spacing, and in-plane acceleration (GRAPPA-generalized autocalibrating partially parallel acquisitions) factor of 2, with a total acquisition time of approximate time of 7 minutes. The blood-oxygen-level-dependent (BOLD) fMRI responses were measured as a function of time using T2*-weighted single-shot multiband accelerated (factor of 7) gradient-echo echo-planar image (EPI) sequence [TE/TR=35/1000 ms, 2.1 isotropic mm resolution, 70 axial slices without gaps for the whole brain (147mm) coverage, FOV 206 × 181 mm, matrix size 96 × 84, 60°-flip angle (approximately Ernst angle), blipped CAIPIRINHA (Controlled Aliasing in Parallel Imaging Results in Higher Acceleration) phase-encoding shift=FOV/3, 1860 kHz/Pixel bandwidth with ramp sampling, echo spacing 0.68 ms, and echo train length 84 ms]. The cue-reactivity task was administrated over three, approximately 6 min 54 s, functional runs. The 2-hour scan included additional structural and functional procedures reported elsewhere.(11–16)

Raw BOLD-fMRI data in DICOM (Digital Imaging and Communications in Medicine) format were first converted to NIFTI (Neuroimaging Informatics Technology Initiative) via dcm2nii(17) and HeuDiConv (<https://github.com/nipy/heudiconv>) and adapted to the BIDS (Brain Imaging Data Structure) format(18) and then preprocessed via fMRIprep pipeline (version 20.2.1).(19, 20) The structural images were Intensity-normalized and skull stripped with ANTs (Advanced Normalization Tools).(21, 22) Volume-based spatial normalization through nonlinear registration into ICBM (International Consortium for Brain Mapping) 152 Nonlinear Asymmetrical template was performed with ANTs.(21, 23) Brain tissue was segmented into the cerebrospinal fluid, white matter, and gray matter through FSL’s [Functional MRI of the Brain (FMRIB) Software Library] FAST (FMRIB Automated Segmentation Tool).(24) Susceptibility distortion correction using echo-planar field maps acquired in opposing phase-encoding directions was applied to the functional images via 3dQwrap in AFNI (Analysis of Functional Neuro Images).(25) Motion artifacts were estimated and corrected for the functional images via FSL’s MCFLIRT (FMRIB Linear Image Registration Tool with motion correction).(26) Motion- and distortion- corrected images were then co-registered to participants’ structural T1w images with the boundary-based registration with 9 degrees of freedom using FSL’S FLIRT,(23, 27) and normalized to ICBM 152 nonlinear asymmetrical template.(23) Preprocessed functional images were visually inspected and those deemed poorly-registered underwent Freesurfer’s recon-all(28) prior to repeating fMRIprep’s preprocessing pipeline (n=6 for men with HUD, n=2 for women with HUD, n=1 for women with CUD).

In addition to fMRIprep, we identified volumes with spikes in translation and rotation parameters using a typical boxplot threshold (75th percentile + 1.5 times the interquartile range) in relation to a reference image volume using FSL’s fsl_motion_outlier. On average, we regressed out 5.49% of the total volumes in each run (range: 1.24%-13.15%) for the first fMRI (for sex differences effects) and 5.38% of the total volumes in each run (range: 2.65%-10.67%) for the second fMRI (for menstrual cycle effects within the 16 women). There were no significant differences in the percentage of motion outliers between men (mean = 5.75% ± 2.56%) and women (mean = 5.10% ± 2.18%; W = 683, p = 0.33) in the first fMRI. Additionally, the frame-wise displacement showed no significant differences between men (mean = 0.30 ± 0.14) and women (mean = 0.31 ± 0.11; W = 901.5, p = 0.26) in the first fMRI. Among the 16 women who underwent two scans, there were no significant differences in the percentage of motion outliers between the first (mean = 4.34% ± 1.92%) and second MRI (mean = 5.34% ± 2.17%; t = 0.21, p = 0.85). Similarly, the frame-wise displacement did not significantly differ between the first MRI (mean = 0.30 ± 0.11) and the second MRI (mean = 0.34 ± 0.19; t = 1.26, p = 0.23). A high-pass filter (100 s cutoff) was applied to the functional data to ignore scanner drift. Lastly, the preprocessed data were spatially smoothed with a Gaussian kernel (5-mm full-width at half maximum) to improve signal to noise ratio.

**General linear model**

The GLM regressors included three “look” events, one “reappraise” event, and one “savor” event, sampled from the onset of the corresponding trials (16-second length, convolved with a double gamma hemodynamic response function). Translation and rotation parameters for motion (x, y, and z for each) and motion outlier time points were included as “nuisance” regressors. Fixation events were not modeled, contributing to baseline. The first-level analysis included the drug cue reactivity contrasts (look drug>look neutral or look food), reappraisal contrasts (reappraise drug>look drug or savor food), and savoring contrasts (savor food>look drug or savor food>look food). Sex differences analyses were conducted with separate one-tailed t-test comparing men and women on each contrast of interest. Paired t-test (one-tailed) was used for examining menstrual cycle effects (i.e., follicular vs. luteal phase) within the women. For within women correlation analyses, we first conducted within-subject subtraction of covariates related to drug cravings and serum hormonal levels (delta directions: follicular minus luteal phase for estradiol and drug craving, luteal minus follicular phase for progesterone and scaled progesterone/estradiol ratio) and subject-level statistical mapping for each contrast of interest between the menstrual cycle phases (Δ) before running correlations between corresponding (same delta [Δ] direction) Δbrain activation and Δcovariates.

**Anatomical mask**

Guided by results from our previous study,(29, 30) we created a fronto-striatal anatomical mask. The resampled and binarized anatomical mask derived from the 25% thresholded Harvard-oxford atlas which encompassed regions including the supplementary motor area, middle frontal gyrus, inferior frontal gyrus, anterior cingulate gyrus, paracingulate gyrus, superior frontal gyrus, frontal pole, frontal medial cortex, subcallosal cortex, orbitofrontal cortex, insula, and striatum (Figure S11). In addition to whole-brain analyses, the anatomical mask was used to restrict search to our a priori regions of interest.

**Ovarian hormones**

Objective blood ovarian hormone testing used chemiluminescence microparticle immunoassay (Abbott Architect; CPT Code: 82670 for Estradiol, 84144 for Progesterone) with sensitivity level of 24.00 pg/ml for estradiol and 0.5 ng/ml for progesterone, conducted at the Mount Sinai Hospital Center for Clinical Laboratories. Three estradiol samples were processed at LABCORP (Raritan, NJ) with an electrochemiluminescence immunoassay (ECLIA, Roche) with sensitivity level of 5.0 pg/ml. The assay sensitivity levels for estradiol and progesterone were used as the data point when the sample’s reading fell below these thresholds. This procedure accounted for a total of 14 samples across 9 participants (5 during the luteal phase and 9 during the follicular phase) which had progesterone readings as 0.5 ng/ml; and five of these 14 samples across 3 participants (3 during the luteal phase and 2 during the follicular phase) which had estradiol readings as 24.00 pg/ml (Figure S5). Except for three sessions (±1 day), blood drawings for these samples were completed on the same day of the MRI procedure (missing estradiol/progesterone readings: n=2/3).

Exploratory correlations showed a trend for Δ (between menstrual phase changes) estradiol level to be positively correlated with Δtask-induced drug craving, such that the higher the follicular>luteal estradiol, the more the respective drug craving (r=0.48, p=0.08). This finding aligns with other studies showing that estradiol enhances drug-seeking behavior.(31, 32) We additionally found Δprogesterone and Δprogesterone/estradiol ratio to be positively correlated with Δcue-induced food craving, such that the higher the luteal>follicular progesterone and progesterone/estradiol ratio, the higher the respective food craving (r=0.83, p=0.0015; r=0.82, p=0.0021). These associations with higher Δcue-induced food craving suggest that progesterone might exert its protective effects via increasing craving/wanting of alternative rewards (reinforcers).

**Supplementary analyses for sex differences and menstrual cycle effects**

In the analyses with iHUD only, we found similar sex differences, at a trend level, in drug cue-reactivity where women showed higher mPFC activity than men (anatomically masked; Z>2.56 and p<0.05 for look drug>look food with a cluster size of 231 voxels, peak p<0.001, peak Z=4.25; Z>2.3 and p<0.05 for look drug>look neutral with a cluster size of 270 voxels, peak p<0.001, peak Z=3.47) and in drug reappraisal activity (>look drug) where men exhibited higher frontal eye field (FEF)/dorsolateral PFC (dlPFC) activity than women (anatomically masked; Z>2.56 and p<0.05, with a cluster size of 129 voxels, peak p=0.006, peak Z=3.58). After removing the 7 men and 2 women with current CUD (mentioned above), we still saw the same trend for higher mPFC drug cue-reactivity (anatomically masked; Z>1.65 and p<0.05 for look drug>look food) in the women and higher dlPFC drug reappraisal (anatomically masked; Z>2.56 and p<0.05) in men.

In the analyses comparing iCUD and iHUD within women, we found no significant differences overlapped with the sex differences results reported in the main text. Instead, we found higher drug cue-reactivity in the inferior temporal gyrus/lateral occipital cortex (>look food) in iHUD and in the precentral gyrus (>look neutral) in iCUD. No significant differences in drug reappraisal were found. After removing the 2 iHUD with current CUD and 3 iCUD with current OUD (mentioned above), we found no significant differences between iCUD and iHUD within women across drug cue-reactivity and drug reappraisal.

When comparing men and women in follicular phase (n=6 for MRI1 and n=10 for MRI2), we found higher drug cue-reactivity in the middle frontal gyrus, precuneus, lateral occipital cortex, frontal pole, and middle temporal gyrus (>look food) and intracalcarine cortex (>look neutral) in women. Compared to men, we also found higher drug cue-reactivity in women in the luteal phase (n=10 for MRI1 and n=6 for MRI2) in the middle frontal gyrus/frontal pole (>look food) and occipital pole (>look neutral). No significant drug reappraisal sex differences were found in related analyses.

**Drug cue-reactivity and drug reappraisal results outside our main cortico-striatal regions of interest**

*Sex differences*

In addition to the medial prefrontal cortex (mPFC), we also found higher drug cue-reactivity (>look food) in the precuneus in women compared to men (Table 2). The greater the drug cue-reactivity (>look food) in the posterior cingulate cortex (PCC)/retrosplenial cortex the more the task-induced drug craving in women compared to men (Table 2). Drug reappraisal activity (>look drug) in the thalamus was more negatively correlated with task-induced drug craving in women compared to men (Table 2). Overall, both the precuneus/PCC and thalamus showed patterns similar to those observed in the ventromedial PFC (vmPFC) as reported in the main text (higher drug cue reactivity in women as associated with increased drug craving and higher reappraisal activity as associated with decreased drug craving).

*Menstrual cycle and hormonal effects (and correlations with Δdrug craving)*

Menstrual phase differences

Compared to the follicular phase, the luteal phase showed higher drug cue-reactivity (>look neutral) in the supplementary motor area, in a pattern opposite to results reported in the main text (Table 2). All other results were consistent with those reported in the main text. That is, in addition to the anterior PFC (aPFC) reported in the main text, the luteal phase also showed higher drug reappraisal (>look drug) activity in other regions of the default mode (i.e., middle/superior temporal gyrus) and visual network (i.e., intracalcarine cortex) and others (Table 2).

Correlations with Δhormones

In addition to the vmPFC reported in the main text, the Δdrug cue-reactivity (>look food) was positively correlated with Δestradiol in other default mode network regions such as the PCC and inferior parietal lobule/angular gyrus and visual areas (Table 2). In a pattern different than reported in the main results, the drug cue-reactivity (>look neutral or food) was positively correlated with Δprogesterone in the dorsal attention and somatomotor networks (including the intraparietal sulcus/supramarginal gyrus, superior parietal lobule, and central opercular cortex) and Δprogesterone/estradiol ratio in the dorsal and ventral attention networks (i.e., precuneus and supramarginal gyrus/temporoparietal junction) and visual areas (Table 2). In contrast, and in addition to the vmPFC, FEF/aPFC, and striatum reported in the main text, Δdrug reappraisal (>look drug) was also negatively correlated with Δestradiol in additional default mode network regions inclusive of the precuneus/PCC, inferior parietal lobule/angular gyrus and other mainly visual areas (Table 2).

Correlations with Δdrug craving

In addition to the inferior frontal gyrus (IFG), Δdrug cue-reactivity (>look neutral) in the thalamus and temporal pole also positively correlated with Δcue-induced drug craving (Table 2).

In summary, with the exception of higher drug cue-reactivity in the luteal phase and its positive correlations with progesterone and progesterone/estradiol ratio, most of the more posterior regions like the supplementary motor area, PCC, and inferior/superior parietal lobule (including angular gyrus and supramarginal gyrus) and sub-cortical regions like the thalamus showed patterns of menstrual cycle/hormonal effects and correlations with drug craving similar to those reported in the main text. Particularly, the PCC (the posterior midline node of the default mode network) demonstrated a similar pattern to the vmPFC in its association with Δestradiol, suggesting that estradiol may simultaneously influence self-directed processing in the PCC,(33) relaying internally-directed information to the vmPFC during drug cue processing(34) as part of its potential neural mechanism underlying drug use vulnerability/resilience.

**Additional results related to food savoring (vs. reappraise drug, look food, and look drug) for completeness**

Drug reappraisal (>savor food) activity in the lingual gyrus was more positively correlated with task-induced drug craving in women than men (Table S1). This activity was higher in the aPFC and insula in addition to other mainly somatomotor areas (e.g., supplementary motor area) during the luteal than the follicular phase within women (Figure S6A; Table S1). Higher insula activity in the luteal phase implicates interoceptive functions(35) such as self-awareness and insight into illness(36) during downregulation of drug cues as directly compared to upregulation of food cues. The same drug reappraisal activity (>savor food) was also negatively correlated Δestradiol in the FEF/aPFC (anatomically masked), caudate (Figure S6B), and other areas (i.e., inferior parietal lobule/angular gyrus, precuneus and visual areas; Table S1) while it was positively correlated with Δprogesterone/estradiol ratio in the dlPFC/IFG (Figure S6C) and other ventral and dorsal attention network regions (i.e., supramarginal gyrus/temporoparietal junction and middle temporal gyrus; Table S1). These results suggest that, compared to food savoring, drug reappraisal activity still exhibited similar cortico-striatal activity patterns to those observed during comparisons to drug cue-reactivity (i.e., drug reappraisal>look drug).

Food savoring (>look food) was higher during the follicular than luteal phase in the dlPFC (Figure S6A), lateral occipital cortex, and precentral gyrus (Table S1). Between menstrual phases, Δfood savoring (>look food) was negatively correlated with Δcue-induced drug craving in the superior parietal lobule (Table S1), such that the higher the Δfood savoring, the lower the Δdrug craving. We also found Δfood savoring to negatively correlated with Δprogesterone/estradiol ratio mostly in visual areas in addition to the intraparietal sulcus/supramarginal gyrus in the dorsal attention network, such that the higher the Δfood savoring, the lower the Δprogesterone/estradiol ratio. Taken together, food savoring may be effortful during the follicular phase and associated with lower craving and progesterone between menstrual phases.

Lastly, we tested effects of food savoring in regulation of drug cue-reactivity and found that, between menstrual phases, Δfood savoring (>look drug) was negatively correlated with Δprogesterone/estradiol ratio in the dlPFC and putamen (Figure S6C) as well as thalamus, FEF, aPFC, superior parietal lobule, and lateral occipital cortex across the dorsal attention, frontoparietal control, and visual networks (Table S1). These findings are consistent with those reported in the above paragraph (negative association between Δfood savoring and Δprogesterone level). A potential explanation is that when progesterone is higher, which is associated with higher food craving (reported above), food savoring becomes more efficient with lower (less effortful) cortico-striatal reactivity.

References

1.	Sheehan DV, Lecrubier Y, Sheehan KH, et al.: The Mini-International Neuropsychiatric Interview (M.I.N.I.): the development and validation of a structured diagnostic psychiatric interview for DSM-IV and ICD-10. J Clin Psychiatry 1998; 59 Suppl 20:22-33;quiz 34-57

2.	McLellan AT, Kushner H, Metzger D, et al.: The fifth edition of the addiction severity index. Journal of Substance Abuse Treatment 1992; 9:199–213

3.	Gossop M, Griffiths P, Powis B, et al.: Severity of dependence and route of administration of heroin, cocaine and amphetamines. British Journal of Addiction 1992; 87:1527–1536

4.	Tiffany ST, Singleton E, Haertzen CA, et al.: The development of a cocaine craving questionnaire. Drug and Alcohol Dependence 1993; 34:19–28

5.	Heinz AJ, Epstein DH, Schroeder JR, et al.: Heroin and cocaine craving and use during treatment: measurement validation and potential relationships. Journal of Substance Abuse Treatment 2006; 31:355–364

6.	Gossop M: The development of a short opiate withdrawal scale (SOWS). Addictive Behaviors 1990; 15:487–490

7.	Kampman KM, Volpicelli JR, McGinnis DE, et al.: Reliability and validity of the Cocaine Selective Severity Assessment. Addictive Behaviors 1998; 23:449–461

8.	Parvaz MA, Malaker P, Zilverstand A, et al.: Attention bias modification in drug addiction: Enhancing control of subsequent habits [Internet]. PNAS 2021; 118[cited 2022 Jan 17] Available from: https://www.pnas.org/content/118/23/e2012941118

9.	Froeliger B, Mathew AR, McConnell PA, et al.: Restructuring Reward Mechanisms in Nicotine Addiction: A Pilot fMRI Study of Mindfulness-Oriented Recovery Enhancement for Cigarette Smokers. Evidence-Based Complementary and Alternative Medicine 2017; 2017:e7018014

10.	Glasser MF, Sotiropoulos SN, Wilson JA, et al.: The minimal preprocessing pipelines for the Human Connectome Project. NeuroImage 2013; 80:105–124

11.	King SG, Gaudreault P-O, Malaker P, et al.: Prefrontal-habenular microstructural impairments in human cocaine and heroin addiction. Neuron 2022; 110:3820-3832.e4

12.	Ceceli AO, Huang Y, Gaudreault P-O, et al.: Recovery of anterior prefrontal cortex inhibitory control after 15 weeks of inpatient treatment in heroin use disorder. Nat Mental Health 2024; 1–9

13.	Kronberg G, Ceceli AO, Huang Y, et al.: Naturalistic drug cue reactivity in heroin use disorder: orbitofrontal synchronization as a marker of craving and recovery. medRxiv 2024; 2023.11.02.23297937

14.	Gaudreault P-O, King SG, Malaker P, et al.: Whole-brain white matter abnormalities in human cocaine and heroin use disorders: association with craving, recency, and cumulative use. Mol Psychiatry 2023; 28:780–791

15.	Ceceli AO, Huang Y, Kronberg G, et al.: Common and distinct fronto-striatal volumetric changes in heroin and cocaine use disorders. Brain 2023; 146:1662–1671

16.	Ceceli AO, King SG, McClain N, et al.: The Neural Signature of Impaired Inhibitory Control in Individuals with Heroin Use Disorder. J Neurosci 2023; 43:173–182

17.	Li X, Morgan PS, Ashburner J, et al.: The first step for neuroimaging data analysis: DICOM to NIfTI conversion. Journal of Neuroscience Methods 2016; 264:47–56

18.	Gorgolewski KJ, Auer T, Calhoun VD, et al.: The brain imaging data structure, a format for organizing and describing outputs of neuroimaging experiments. Scientific Data 2016; 3:1–9

19.	Esteban O, Markiewicz CJ, Blair RW, et al.: fMRIPrep: a robust preprocessing pipeline for functional MRI. Nat Methods 2019; 16:111–116

20.	Gorgolewski K, Burns C, Madison C, et al.: Nipype: A Flexible, Lightweight and Extensible Neuroimaging Data Processing Framework in Python. Frontiers in Neuroinformatics 2011; 5:13

21.	Avants BB, Epstein CL, Grossman M, et al.: Symmetric diffeomorphic image registration with cross-correlation: Evaluating automated labeling of elderly and neurodegenerative brain. Medical Image Analysis 2008; 12:26–41

22.	Tustison NJ, Avants BB, Cook PA, et al.: N4ITK: improved N3 bias correction. IEEE Trans Med Imaging 2010; 29:1310–1320

23.	Fonov V, Evans A, McKinstry R, et al.: Unbiased nonlinear average age-appropriate brain templates from birth to adulthood. NeuroImage 2009; 47:S102

24.	Zhang Y, Brady M, Smith S: Segmentation of brain MR images through a hidden Markov random field model and the expectation-maximization algorithm. IEEE Transactions on Medical Imaging 2001; 20:45–57

25.	Cox RW: AFNI: Software for Analysis and Visualization of Functional Magnetic Resonance Neuroimages. Computers and Biomedical Research 1996; 29:162–173

26.	Jenkinson M, Bannister P, Brady M, et al.: Improved optimization for the robust and accurate linear registration and motion correction of brain images. Neuroimage 2002; 17:825–841

27.	Jenkinson M, Smith S: A global optimisation method for robust affine registration of brain images. Medical Image Analysis 2001; 5:143–156

28.	Dale AM, Fischl B, Sereno MI: Cortical Surface-Based Analysis: I. Segmentation and Surface Reconstruction. NeuroImage 1999; 9:179–194

29.	Huang Y, Ceceli AO, Kronberg G, et al.: Association of Cortico-Striatal Engagement During Cue Reactivity, Reappraisal, and Savoring of Drug and Non-Drug Stimuli With Craving in Heroin Addiction. AJP 2024; 181:153–165

30.	Zilverstand A, Huang AS, Alia-Klein N, et al.: Neuroimaging Impaired Response Inhibition and Salience Attribution in Human Drug Addiction: A Systematic Review. Neuron 2018; 98:886–903

31.	Becker JB, McClellan ML, Reed BG: Sex differences, gender and addiction. Journal of Neuroscience Research 2017; 95:136–147

32.	Nicolas C, Zlebnik NE, Farokhnia M, et al.: Sex Differences in Opioid and Psychostimulant Craving and Relapse: A Critical Review. Pharmacol Rev 2022; 74:119–140

33.	Leech R, Sharp DJ: The role of the posterior cingulate cortex in cognition and disease. Brain 2014; 137:12–32

34.	Andrews-Hanna JR, Smallwood J, Spreng RN: The default network and self-generated thought: component processes, dynamic control, and clinical relevance. Annals of the New York Academy of Sciences 2014; 1316:29–52

35.	Naqvi NH, Bechara A: The hidden island of addiction: the insula. Trends in Neurosciences 2009; 32:56–67

36.	Goldstein RZ, Craig AD (Bud), Bechara A, et al.: The Neurocircuitry of Impaired Insight in Drug Addiction. Trends in Cognitive Sciences 2009; 13:372–380

37.	Xia M, Wang J, He Y: BrainNet Viewer: A Network Visualization Tool for Human Brain Connectomics. PLOS ONE 2013; 8:e68910

**Table S1.** **Coordinates for fMRI-BOLD results related to food savoring**

| **Structure** | **Side** | **Voxels** | **Peak Z** | **Peak p** | **X** | **Y** | **Z** | **BA** | **Network** |
| --- | --- | --- | --- | --- | --- | --- | --- | --- | --- |
| **Food savoring related activity** | | | | | | | | | |
| ***Sex effects*** | | | | | | | | | |
| ***Correlations with drug cravings*** | | | | | | | | | |
| ***Women>Men: RD>SF correlation with task-induced (post-pre) craving***  ***driven by positive correlation within women*** | | | | | | | | | |
| Lingual Gyrus | R | 330 | +4.81 | <0.0001 | 9 | -64 | 1 | 18 | Visual |
| ***Menstrual cycle and hormonal effects*** | | | | | | | | | |
| ***Menstrual phase differences*** | | | | | | | | | |
| ***Luteal>Follicular: RD>SF*** | | | | | | | | | |
| Temporal Pole | R | 129 | 4.32 | <0.0001 | 52 | 9 | -22 | 38 | Default |
| Insula ^a^ | R |  | 4.07 | <0.0001 | 39 | 16 | -9 | 13 | Frontoparietal Control |
| Supplementary Motor Area | R | 108 | 4.52 | 0.0002 | 4 | -4 | 66 | 6 | Ventral Attention |
| Temporal Occipital Fusiform Cortex | R | 95 | 4.52 | 0.0004 | 39 | -53 | -13 | 37 | Visual |
| Supplementary Motor Area/Superior Frontal Gyrus | L | 77 | 4.25 | 0.0020 | -17 | -10 | 71 | 6 | Somatomotor |
| Superior Temporal Gyrus (posterior division) | R | 65 | 4.76 | 0.0060 | 67 | -30 | 14 | 22 | Somatomotor |
| Supplementary Motor Area/Precentral Gyrus | R | 65 | 4.45 | 0.0060 | 26 | -23 | 75 | 6 | Somatomotor |
| Middle/Superior Temporal Gyrus (posterior division) | R | 48 | 4.71 | 0.0314 | 54 | -17 | -9 | 22 | Default |
| Anterior Prefrontal Cortex | R | 46 | 4.24 | 0.0385 | 26 | 59 | -7 | 10 | Frontoparietal Control |
| ***Follicular>Luteal: SF>LF*** | | | | | | | | | |
| Precentral Gyrus | R | 83 | 4.67 | 0.0012 | 37 | -25 | 66 | 4 | Somatomotor |
| Dorsolateral Prefrontal Cortex | R | 70 | 4.58 | 0.0039 | 37 | 33 | 43 | 9 | Frontoparietal Control |
| Lateral Occipital Cortex (superior division) | R | 45 | 4.32 | 0.0435 | 28 | -83 | 39 | 7 | Visual |
| ***Correlations with Δhormones*** | | | | | | | | | |
| ***ΔRD>SF negative correlation with ΔE*** | | | | | | | | | |
| Frontal Eye Field/Anterior Prefrontal Cortex ^b^ | R | 33 | -4.65 | 0.0331 | 15 | 44 | 52 | 8 | Default |
| Intracalcarine Cortex | R | 206 | -4.74 | <0.0001 | 11 | -85 | 3 | 17 | Visual |
| Cerebellum Crus II | R | 181 | -4.46 | <0.0001 | 32 | -79 | -41 |  |  |
| Inferior Parietal Lobule (Angular Gyrus) | R | 80 | -4.27 | 0.0016 | 34 | -70 | 35 | 39 | Dorsal Attention |
| Caudate | R | 72 | -4.89 | 0.0032 | 11 | -4 | 18 |  |  |
| Precuneus | L | 65 | -4.15 | 0.0060 | -2 | -74 | 54 | 7 | Frontoparietal Control |
| Cerebellum VI | L | 55 | -4.53 | 0.0157 | -32 | -55 | -30 |  |  |
| Inferior Parietal Lobule (Angular Gyrus) | L | 51 | -4.45 | 0.0233 | -43 | -77 | 37 | 39 | Default |
| Occipital Pole | L | 50 | -4.53 | 0.0258 | -13 | -96 | -9 | 18 | Visual |
| Retrosplenial Cortex | R | 49 | -4.43 | 0.0286 | 2 | -45 | 8 | 30 | Default |
| Cerebellum Crus II | L | 44 | -4.89 | 0.0478 | -17 | -90 | -32 |  |  |
| Frontal Eye Field/Anterior Prefrontal Cortex ^a^ | R | 33 | -4.65 | 0.0331 | 15 | 44 | 52 | 8 | Default |
| Intracalcarine Cortex | R | 206 | -4.74 | <0.0001 | 11 | -85 | 3 | 17 | Visual |
| ***ΔRD>SF positive correlation with ΔP/E ratio*** | | | | | | | | | |
| Supramarginal Gyrus/Temporoparietal Junction | L | 71 | +4.59 | 0.0029 | -64 | -34 | 35 | 40 | Ventral Attention |
| Middle Temporal Gyrus (temporooccipital part) | L | 66 | +4.51 | 0.0046 | -60 | -53 | -5 | 37 | Frontoparietal Control |
| Middle Temporal Gyrus (temporooccipital part) | R | 60 | +4.34 | 0.0083 | 56 | -57 | 1 | 37 | Dorsal Attention |
| Postcentral Gyrus | L | 58 | +4.38 | 0.0101 | -47 | -40 | 60 | 1 | Dorsal Attention |
| Superior Temporal Gyrus (posterior division) | L | 55 | +4.16 | 0.0136 | -51 | -42 | 12 | 22 | Default |
| Precentral Gyrus | R | 48 | +4.37 | 0.0279 | 2 | -30 | 60 | 4 | Somatomotor |
| Inferior Frontal Gyrus/Dorsolateral Prefrontal Cortex | R | 46 | +4.53 | 0.0344 | 39 | 41 | 10 | 46 | Frontoparietal Control |
| ***ΔSF>LF negative correlation with ΔP/E ratio*** | | | | | | | | | |
| Lateral Occipital Cortex (inferior division) | R | 194 | -4.78 | <0.0001 | 52 | -75 | 1 | 19 | Visual |
| Precuneous | L | 64 | -4.60 | 0.0068 | -11 | -57 | 6 | 23 | Visual |
| Intraparietal Sulcus (Supramarginal Gyrus) | R | 64 | -4.20 | 0.0068 | 39 | -42 | 41 | 7 | Dorsal Attention |
| Inferior Temporal Gyrus (temporooccipital part) | R | 61 | -4.39 | 0.0090 | 47 | -60 | -7 | 37 | Visual |
| Temporal Fusiform Cortex | R | 46 | -4.30 | 0.0397 | 32 | -36 | -20 | 37 | Visual |
| ***ΔSF>LD negative correlation with ΔP/E ratio*** | | | | | | | | | |
| Thalamus | L | 135 | -4.64 | <0.0001 | -9 | -12 | 10 |  |  |
| Thalamus | R | 107 | -4.3 | <0.0001 | 17 | -23 | 12 |  |  |
| Dorsolateral Prefrontal Cortex | R | 86 | -4.8 | 0.0006 | 19 | 44 | 31 | 9 | Default |
| Putamen | R | 61 | -4.55 | 0.0063 | 28 | 9 | 8 |  |  |
| Postcentral Gyrus | L | 59 | -4.96 | 0.0077 | -34 | -38 | 64 | 1 | Somatomotor |
| Frontal Eye Field/Anterior Prefrontal Cortex | R | 57 | -4.5 | 0.0095 | 28 | 26 | 52 | 8 | Frontoparietal Control |
| Superior Parietal Lobule | L | 52 | -4.06 | 0.0159 | -11 | -55 | 71 | 7 | Dorsal Attention |
| Lateral Occipital Cortex (inferior division) | R | 48 | -4.24 | 0.0243 | 49 | -70 | 8 | 19 | Visual |
| Anterior Prefrontal Cortex | R | 44 | -4.58 | 0.0374 | 28 | 54 | 10 | 10 | Frontoparietal Control |
| ***Correlations with Δdrug cravings*** | | | | | | | | | |
| ***ΔSF>LF negative correlation with Δcue-induced drug craving*** | | | | | | | | | |
| Superior Parietal lobule | R | 64 | -4.17 | 0.0096 | 32 | -53 | 56 | 7 | Dorsal Attention |

**^a^** Indicates a local maxima coordinate.

**^b^** Indicates results from restricted search using the independent anatomical mask.

Positive and negative Z scores indicate positive and negative correlations respectively.

Network labels for peak coordinates are based on the 7 networks (Yeo et al., 2011) derived from 400 parcellation (Schaefer et al., 2018).

BA=Brodmann Area; Δ=differences between menstrual phases; LD=look drug; LF=look food; RD=reappraise drug; SF=savor food; E=estradiol; P=progesterone; P/E ratio=Progesterone/Estradiol ratio (scaled).

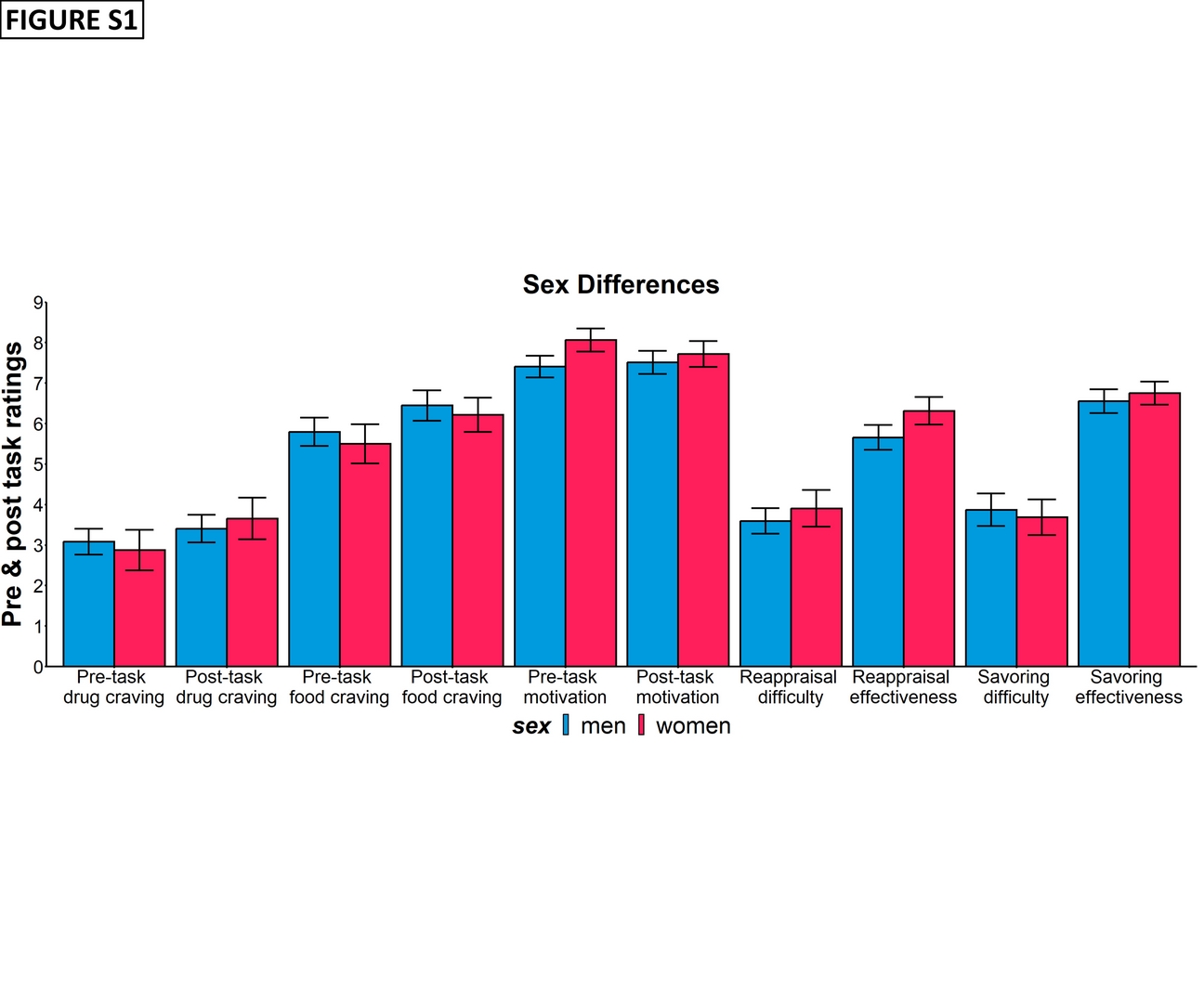

**Figure S1: Bar plots for sex differences of pre- and post-task ratings**

All the error bars denote the mean ± SE.

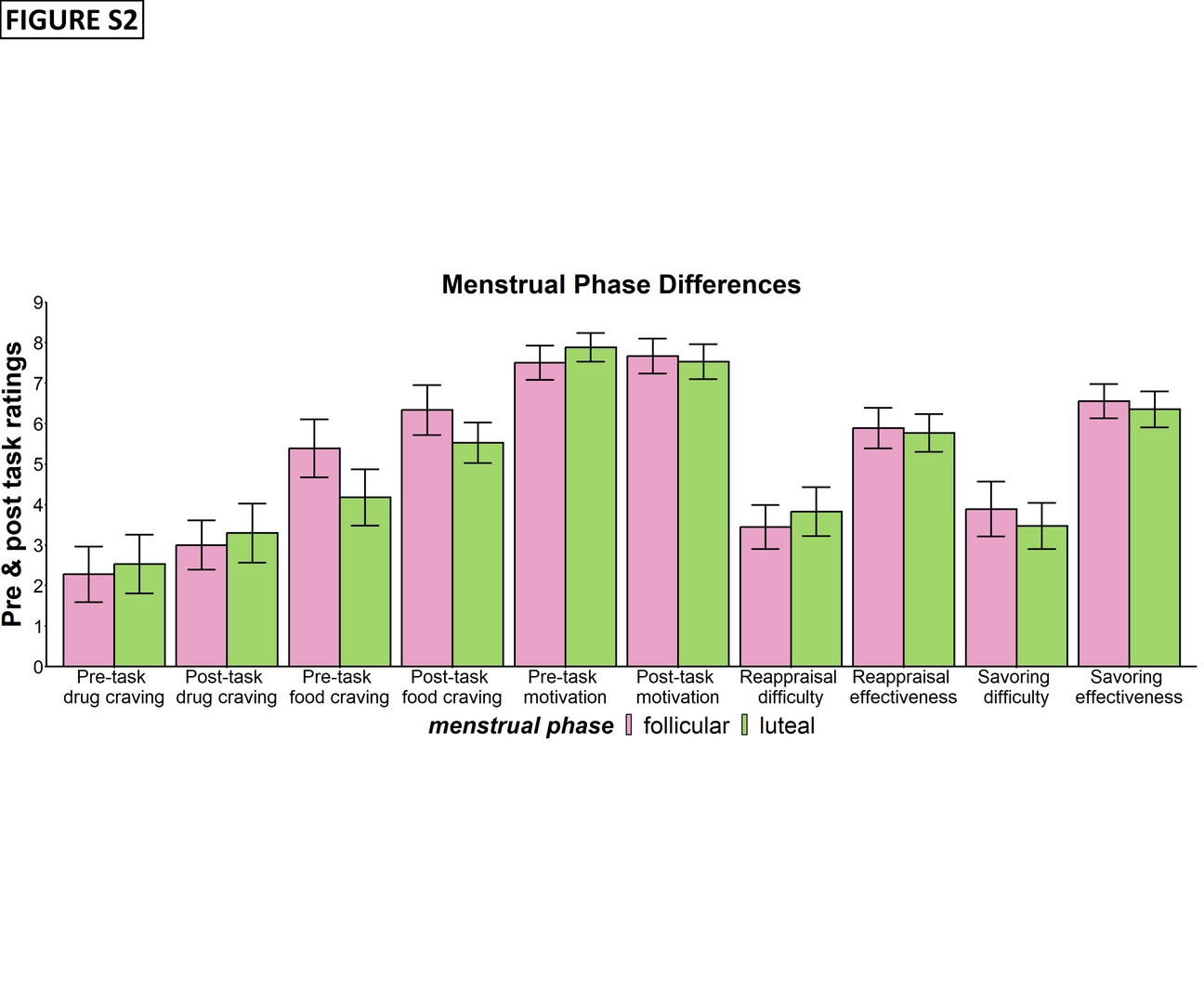

**Figure S2: Bar plots for menstrual phase differences of pre- and post-task ratings**

All the error bars denote the mean ± SE.

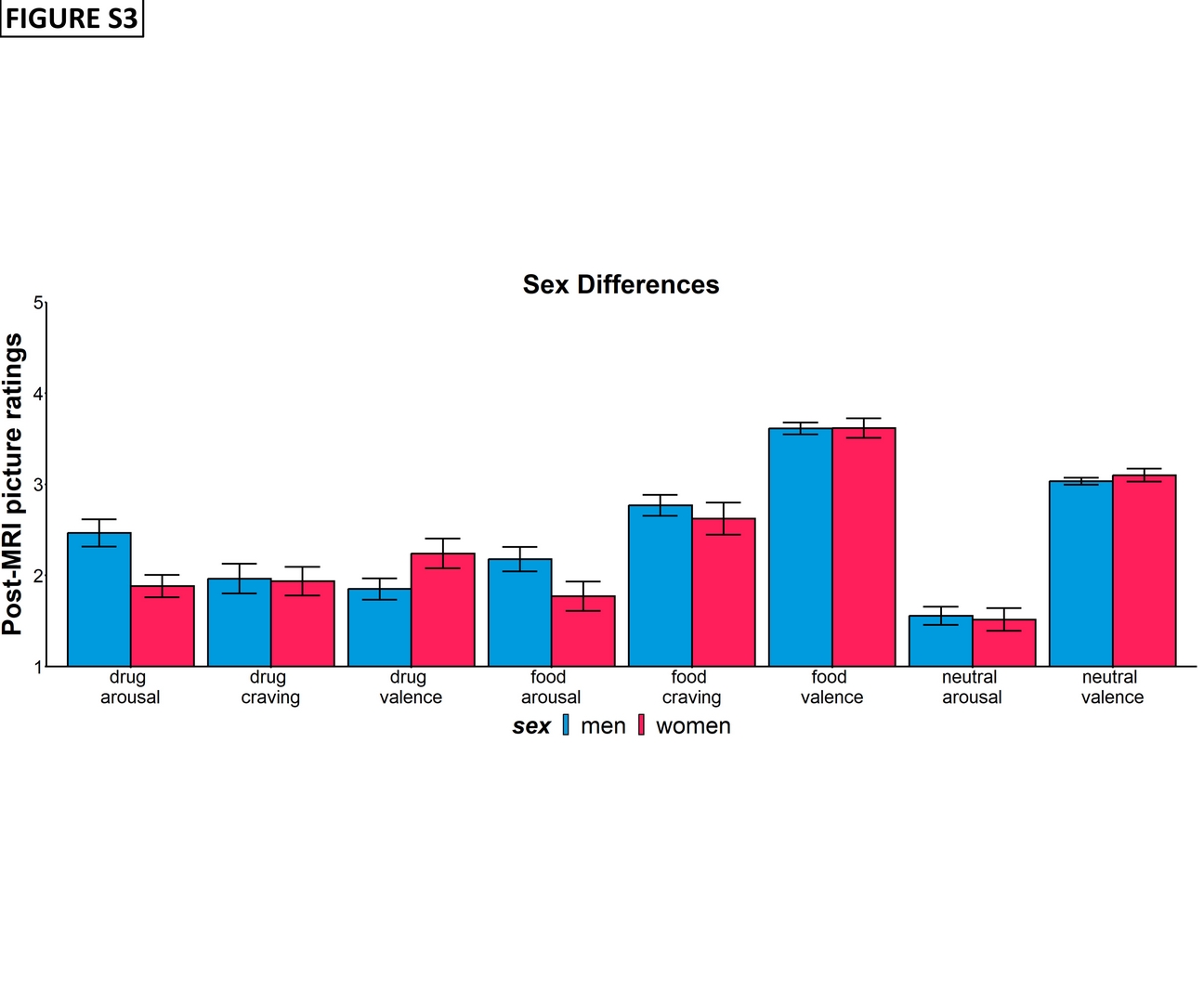

**Figure S3: Bar plots for sex differences of post-MRI picture ratings**

All the error bars denote the mean ± SE.

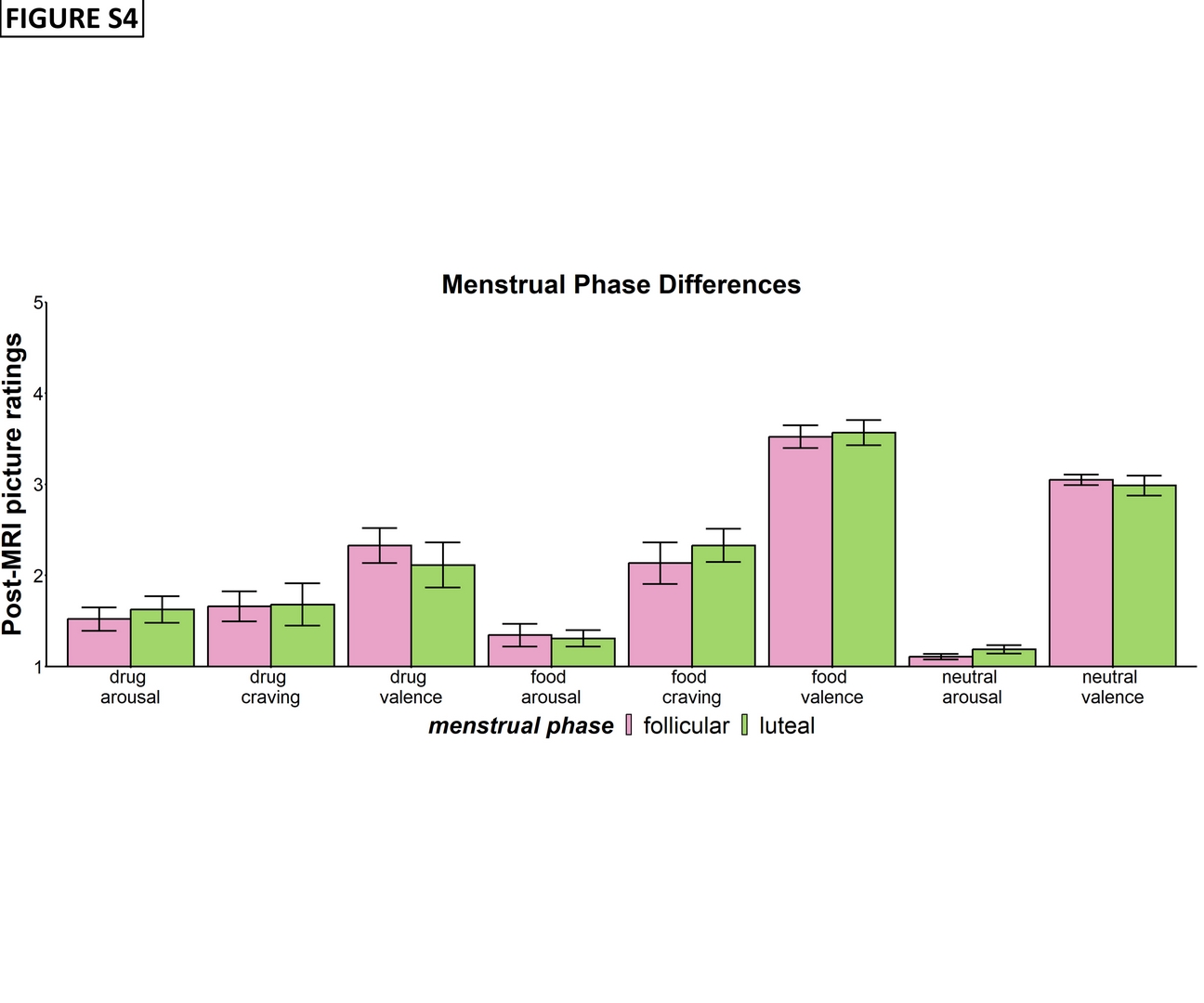

**Figure S4: Bar plots for menstrual phase differences of post-MRI picture ratings**

All the error bars denote the mean ± SE.

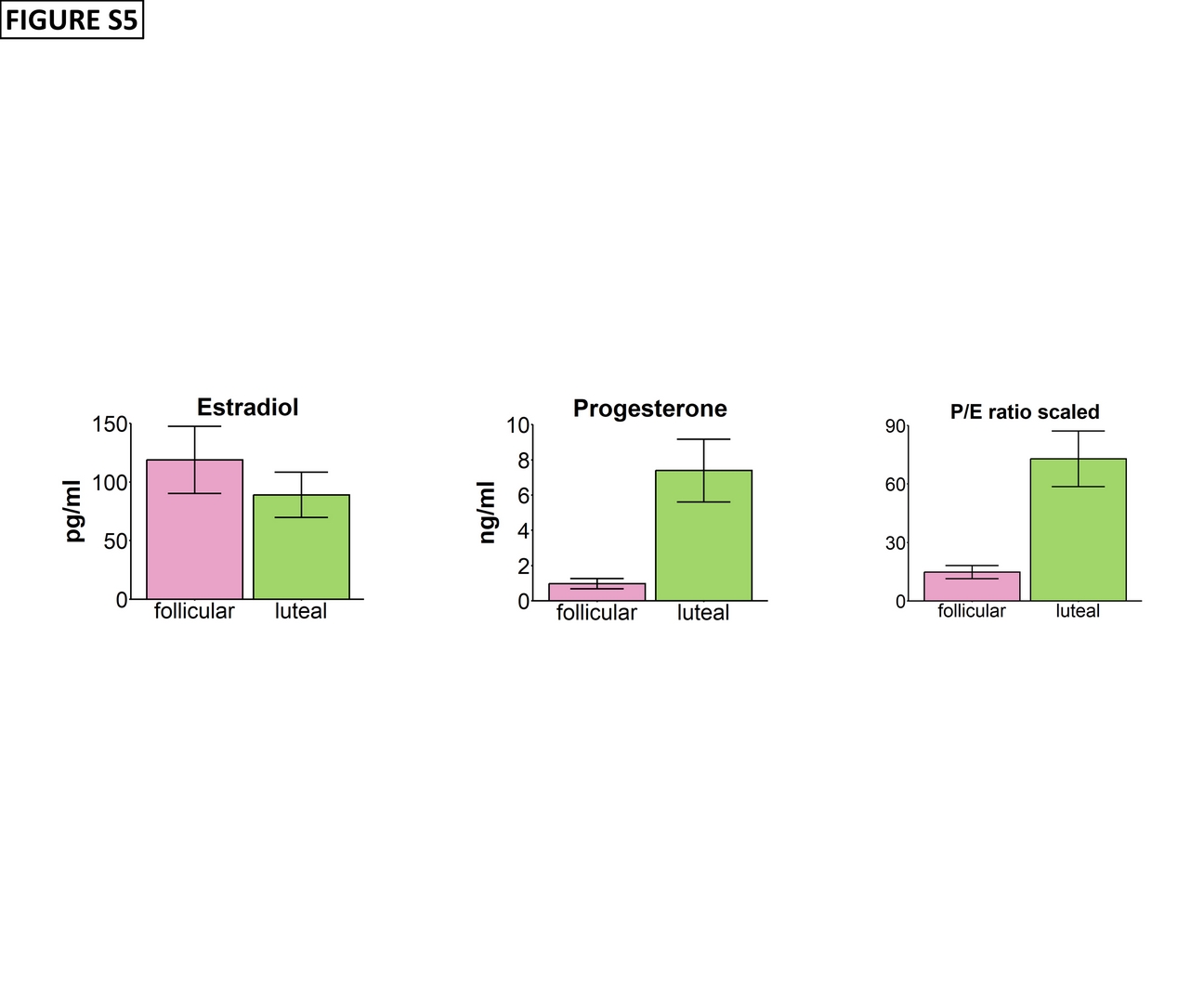

**Figure S5: Within-women ovarian hormone level between follicular and luteal phases**

Estradiol (pg/ml) level plot on the left, progesterone (ng/ml) level plot on the middle and scaled (i.e., progesterone unit was converted to pg/ml before calculating ratio to estradiol individually) and progesterone to estradiol (P/E) ratio plot on the right. All the error bars denote the mean ± SE.

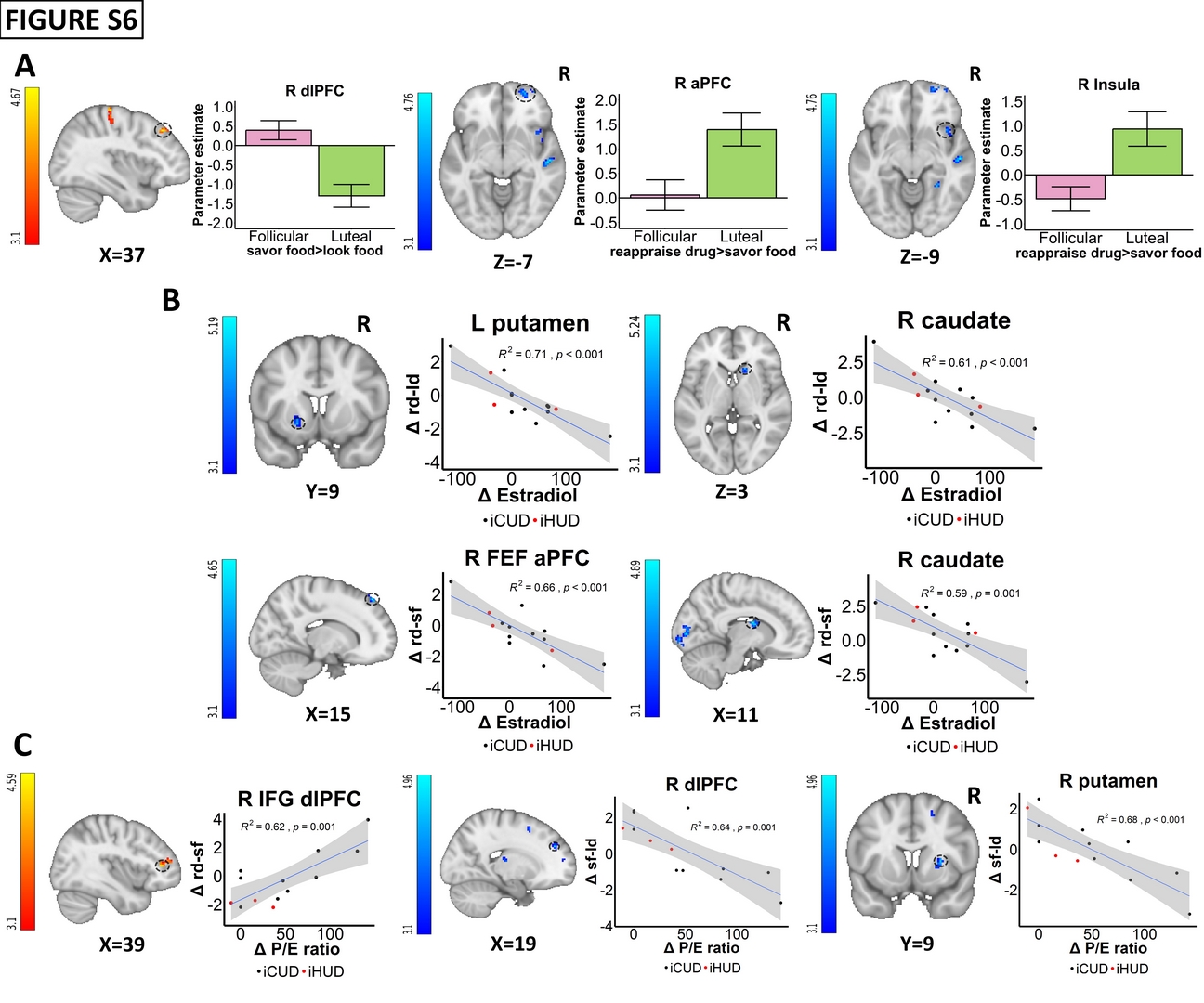

**Figure S6: Supplementary plots for menstrual cycle and hormonal effects**

**A: Menstrual phase differences.** Food savoring (>look food) activity was higher in the dorsolateral prefrontal cortex (dlPFC) during the follicular phase and drug reappraisal (>savor food) activity were higher in the anterior PFC (aPFC) and insula during the luteal phase. **B: Correlations with Δ (between menstrual phase changes) estradiol.** The Δdrug reappraisal (>look drug or savor food) activity were negatively correlated with Δestradiol in the striatum (caudate and putamen) and the frontal eye field (FEF)/aPFC (anatomically masked). **C: Correlations with Δ progesterone to estradiol (P/E) ratio.** Δdrug reappraisal (>savor food) was positively correlated with ΔP/E ratio in the inferior frontal gyrus (IFG)/dlPFC. The Δfood savoring (>look drug; regulation of drug cue-reactivity) activity in dlPFC, and putamen were negatively correlated with ΔP/E ratio. iCUD = individuals with cocaine use disorder; iHUD = individuals with heroin use disorder; rd-ld=reappraise drug>look drug; rd-sf=reappraise drug>savor food; sf-ld=savor food>look drug; L=left; R=right. For visualization purposes, parameter estimates, depicting blood-oxygen-level-dependent signal, were extracted from corresponding FSL zstat images via 3-mm radius masks centered on Montreal Neurological Institute coordinates from peak activity (black dotted line circles represent the approximate peak coordinates). R^2^ and p values were derived from extracted values. All the error bars denote the mean ± SE.

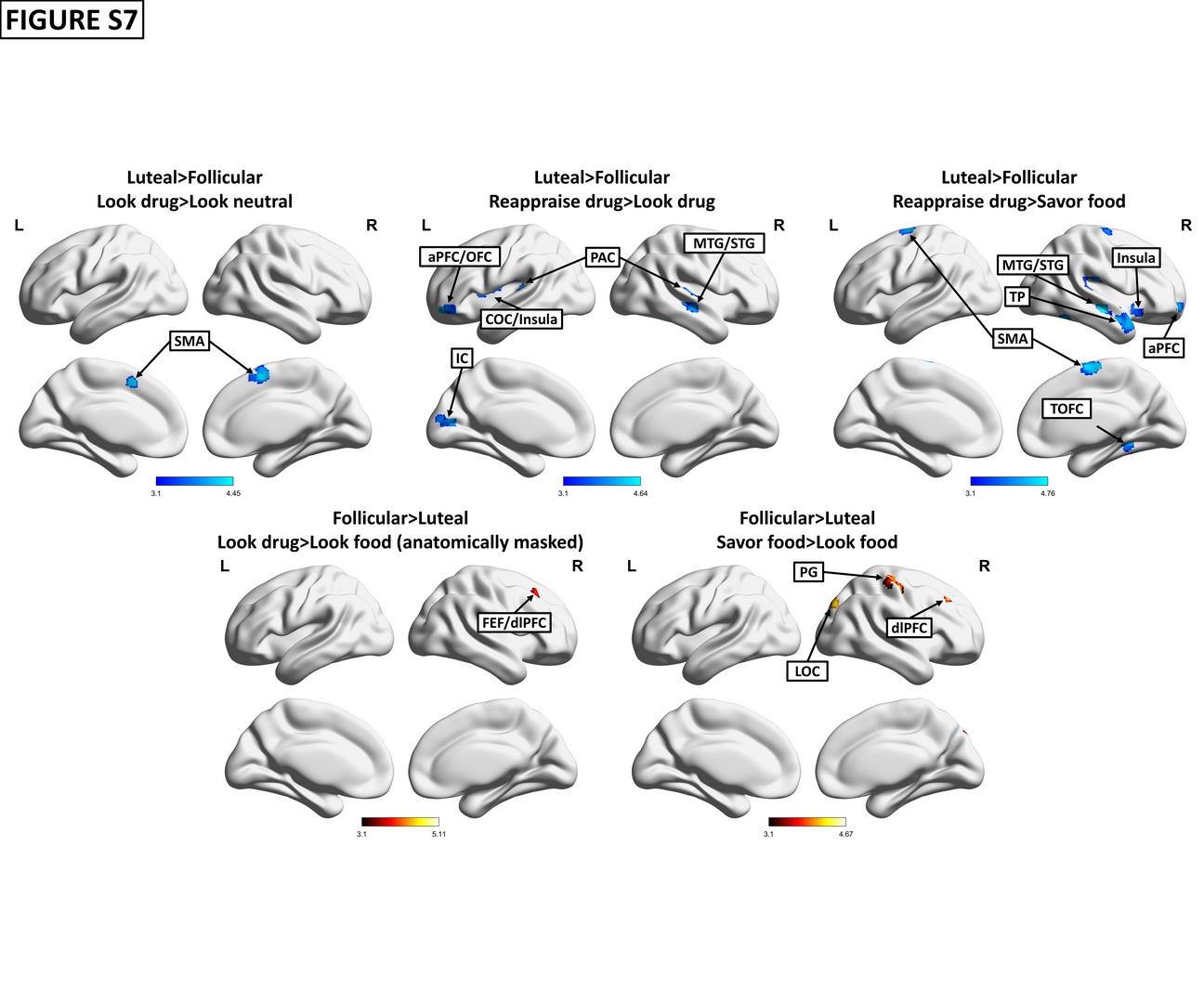

**Figure S7: fMRI BOLD menstrual cycle effects**

The 1^st^ row demonstrates higher activity during the luteal than the follicular phase for drug cue reactivity (look drug>look neutral) and drug reappraisal (reappraise drug>look drug and reappraise drug>savor food contrasts). The 2^nd^ row demonstrated higher activity during the follicular than the luteal phase for drug cue reactivity (look drug>look food, anatomically masked) and food savoring (savor food>look food). SMA = supplementary motor cortex; PG = precentral gyrus; aPFC = anterior prefrontal cortex; OFC = orbitofrontal cortex; MTG/STG = middle/superior temporal gyrus; TP = temporal pole; PAC = primary auditory cortex; COC = central opercular cortex; TOFC = temporal occipital fusiform cortex; IC = intracalcarine cortex; LOC = lateral occipital cortex; FEF = frontal eye field; dlPFC = dorsolateral prefrontal cortex. Visualization was carried out using BrainNet Viewer(37) via the maximum voxel mapping algorithm projecting activations to the cortical surface (smoothed ICBM152 brain).

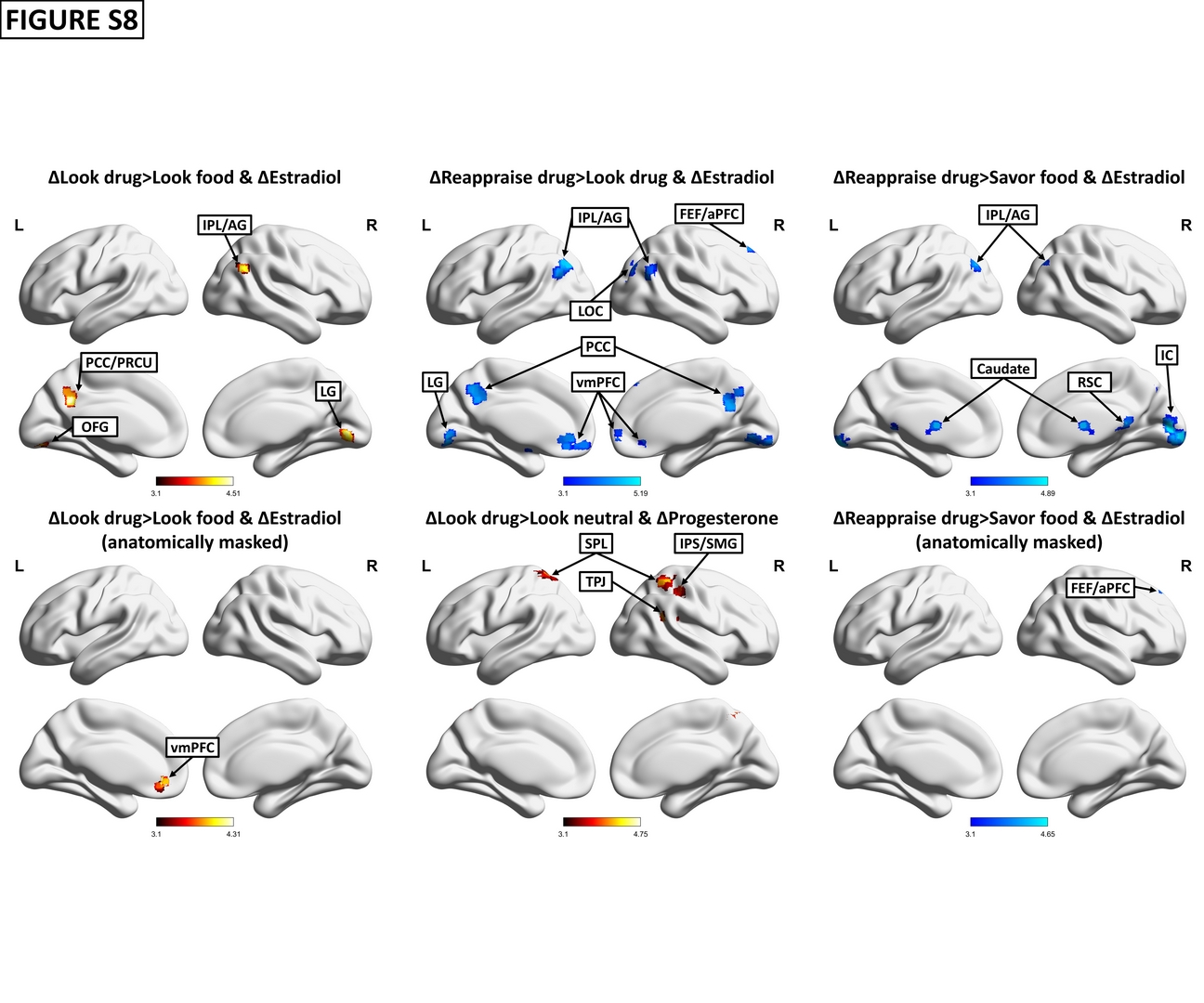

**Figure S8: fMRI BOLD showing correlations with Δ (changes between menstrual phases) estradiol and Δprogesterone**

These figures show the correlations: 1) Δestradiol was positively associated with Δdrug cue-reactivity (>look food; both whole brain and anatomically masked) and negatively associated with Δdrug reappraisal (>look drug or savor food [both whole brain and anatomically masked]); 2) Δprogesterone was positively correlated with Δdrug cue-reactivity (>look neutral). IPL/SPL = inferior/superior parietal lobule; AG = angular gyrus; SMG = supramarginal gyrus; IPS = intraparietal sulcus; TPJ = temporoparietal junction; PCC = posterior cingulate cortex; PRCU = precuneus; RSC = retrosplenial cortex; LG = lingual gyrus; IC = intracalcarine cortex; LOC = lateral occipital cortex; OFG = occipital fusiform gyrus; FEF = frontal eye field; aPFC = anterior prefrontal cortex; vmPFC = ventromedial prefrontal cortex. Visualization was carried out using BrainNet Viewer(37) via the maximum voxel mapping algorithm projecting activations to the cortical surface (smoothed ICBM152 brain).

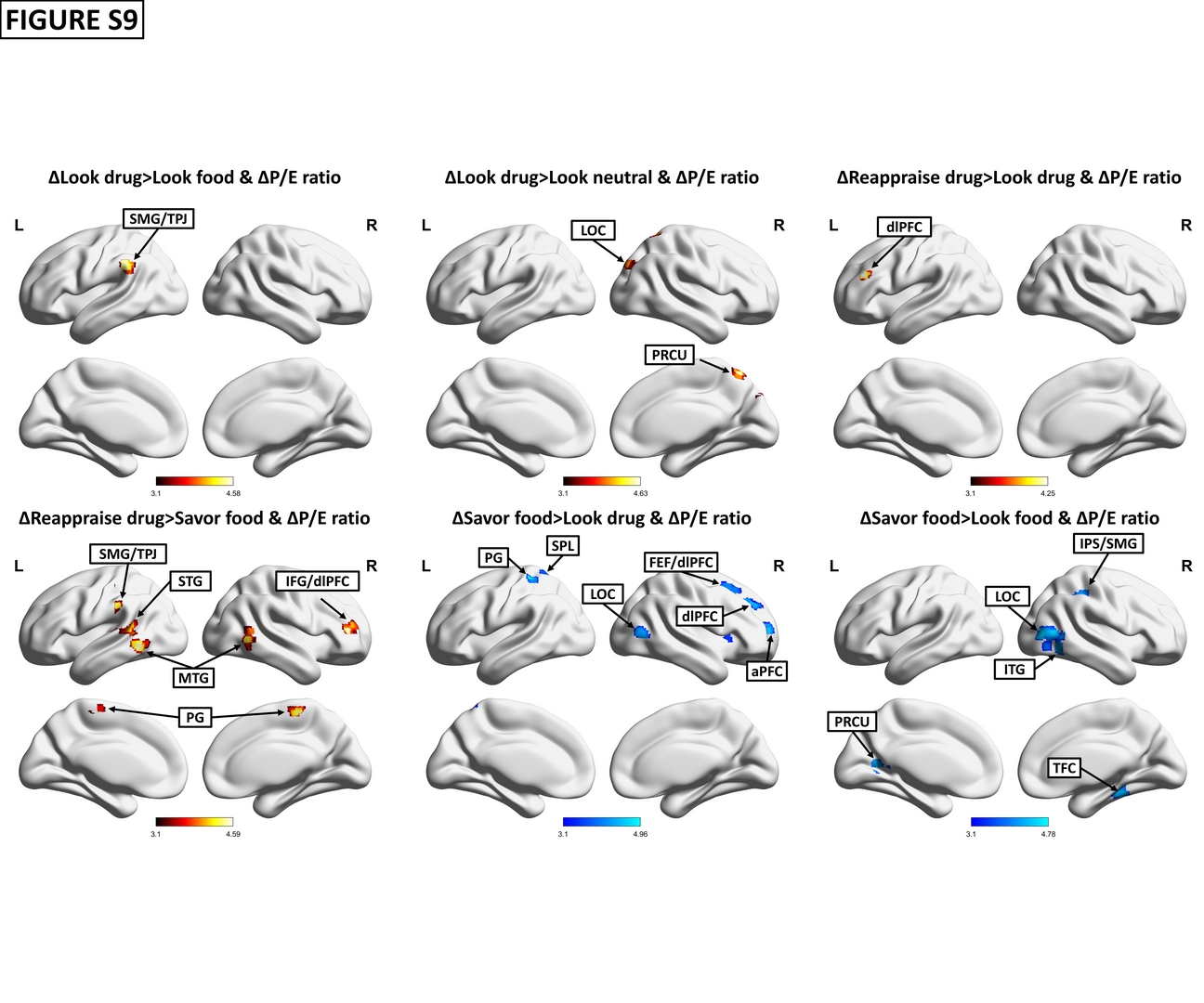

**Figure S9: fMRI BOLD for correlations with Δ (changes between menstrual phases) progesterone to estradiol (P/E) ratio**

ΔP/E ratio were positively correlated with Δdrug cue-reactivity (>look food or look neutral) and Δdrug reappraisal (>look drug or savor food) activity and negatively correlated with Δfood savoring (>look food or look drug [regulation of drug cue-reactivity]) activity. SMG = supramarginal gyrus; TPJ = temporoparietal junction; IPS = intraparietal sulcus; LOC = lateral occipital cortex; dlPFC = dorsolateral prefrontal cortex; IFG = inferior frontal gyrus; PRCU = precuneus; ITG/MTG/STG = inferior/middle/superior temporal gyrus; PG = precentral gyrus; TFC = temporal fusiform cortex. Visualization was carried out using BrainNet Viewer(37) via the maximum voxel mapping algorithm projecting activations to the cortical surface (smoothed ICBM152 brain).

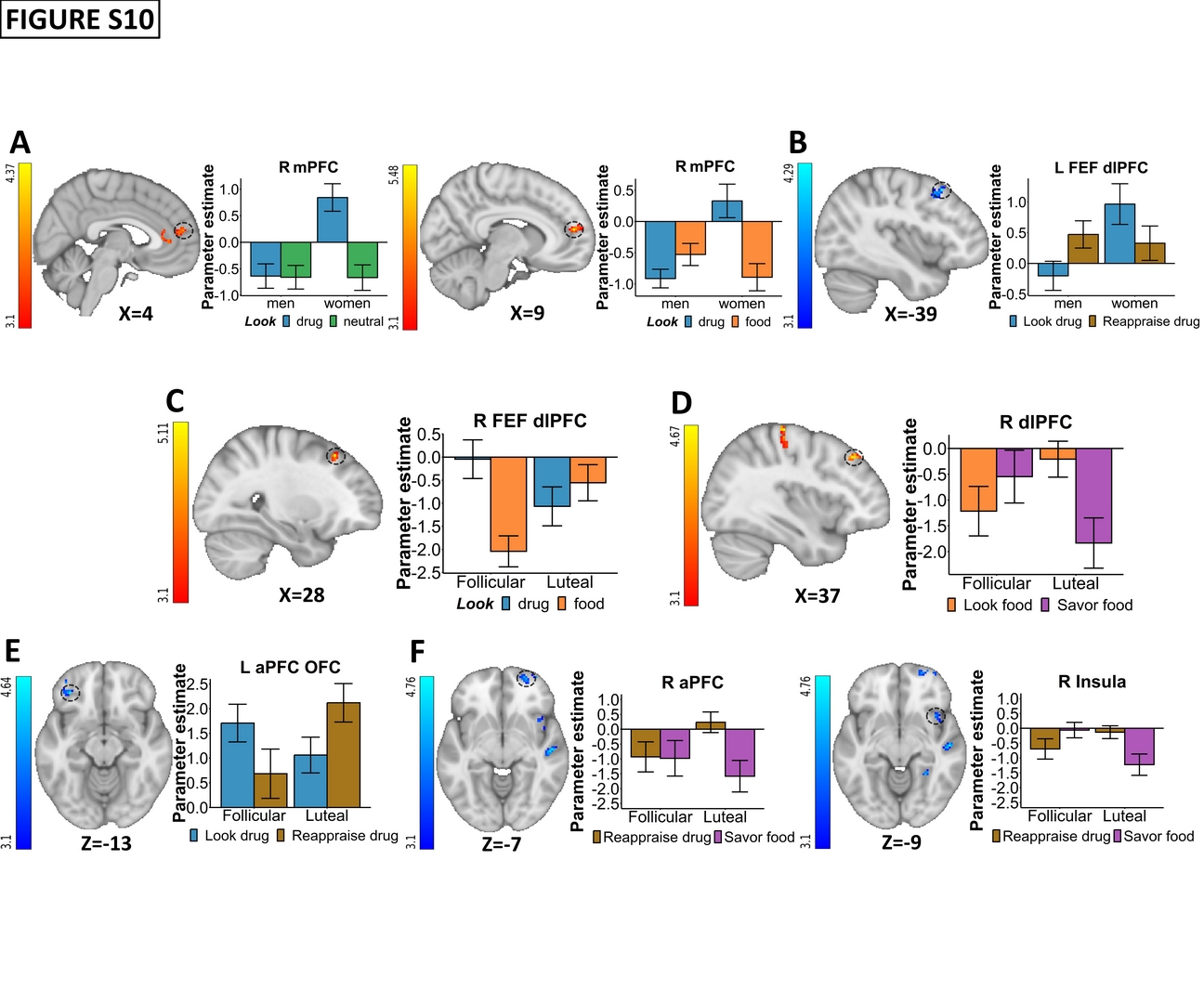

**Figure S10: Condition vs. implicit baseline plots**

A-F corresponding to main Figures 1A, 1B, 2A, S6A (left plot), 3A, and S6A (middle and right plot). For visualization purposes, parameter estimates, depicting the blood-oxygen-level-dependent signal, were extracted from corresponding FSL zstat images via 3-mm radius masks centered on Montreal Neurological Institute coordinates from peak activity (black dotted line circles represent the approximate peak coordinates). All the error bars denote the mean ± SE.

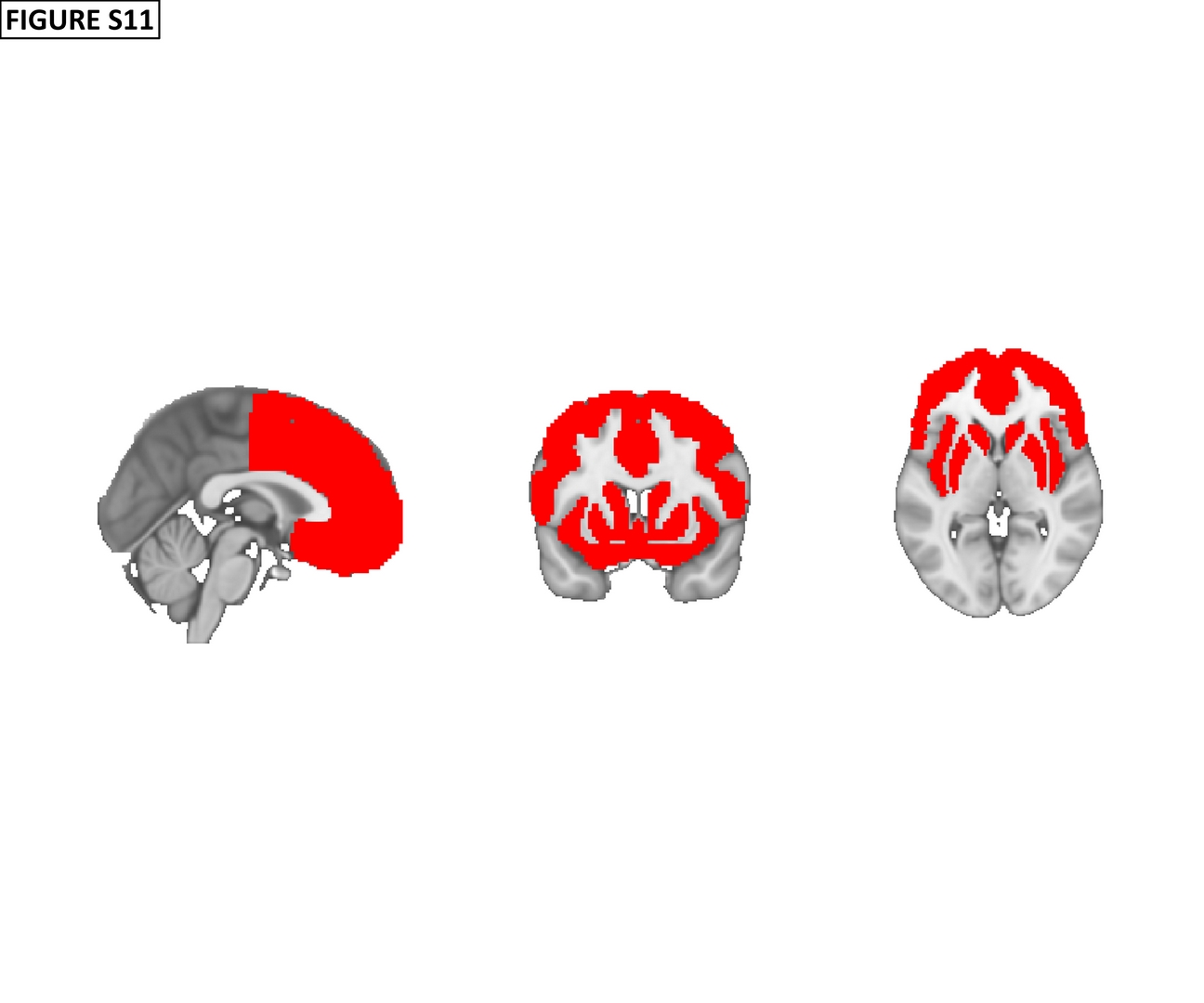

**Figure S11: Independent frontal cortical and striatal anatomical mask**

The resampled and binarized anatomical mask was derived from a 25% thresholded Harvard-oxford atlas including the supplementary motor cortex, frontal pole, frontal medial cortex, paracingulate gyrus, anterior cingulate gyrus, subcallosal cortex, superior frontal gyrus, middle frontal gyrus, orbitofrontal cortex, inferior frontal gyrus, insula, putamen, caudate and nucleus accumbens.
